# School Distress in UK school children: A story dominated by neurodivergence and unmet needs

**DOI:** 10.1101/2022.09.28.22280324

**Authors:** Sophie E. Connolly, Hannah Constable, Sinéad L. Mullally

## Abstract

**Background:** The Covid-19 pandemic has brought into sharp focus a school attendance crisis in many countries, although this likely pre-dates the pandemic. Children and young people (CYP) with school attendance problems (SAPs) often display extreme emotional distress when required to attend school. We term this School Distress (SD). Here we sought to elucidate the characteristics of the CYP struggling to attend school in the UK.

**Methods:** Using a case-control, concurrent embedded mixed-method research design, 947 parents of CYP with experience of SD completed a bespoke online questionnaire (February/March 2022), alongside an aged-matched control group (n=149) and a smaller group of parents who electively home-educate (n=25).

**Results:** In 94.3% of cases, SAPs were underpinned by significant emotional distress, with often harrowing accounts of this distress provided by parents. Whilst the mean age of the CYP in this sample was 11.6 years (StDev 3.1 years), their SD was evident to parents from a much younger age (7.9 years). Notably, 92.1% of CYP currently experiencing SD were described as neurodivergent (ND) and 83.4% as autistic. The Odds Ratio of autistic CYP experiencing SD was 46.61 (95% CI [24.67, 88.07]). Autistic CYP displayed SD at a significantly earlier age, and it was significantly more enduring. Multi-modal sensory processing difficulties and ADHD (amongst other ND conditions) were also commonly associated with SD; with SD CYP having an average of 3.62 NDs (StDev 2.68). In addition, clinically significant anxiety symptomology (92.5%; ASC-ASD-P) and elevated demand avoidance (EDA-8) were also pervasive. Mental health difficulties in the absence of a ND profile were, however, relatively rare (6.17%). Concerningly, despite the striking levels of emotional distress and disability reported by parents, parents also reported a dearth of meaningful support for their CYP at school.

**Conclusion:** Whilst not a story of exclusivity relating solely to autism, SD is a story dominated by complex neurodivergence and a seemingly systemic failure to meet the needs of these CYP in UK schools. Given the disproportionate number of disabled CYP impacted, we ask whether the UK is upholding its’ responsibility to ensure the “right to an education” for all CYP (Human Rights Act 1998).

## Introduction

> A withered boy who was so afraid, hiding from society in the shade, His solitary cries no-one did hear, his confused mind full of fear. His tortured soul locked inside, with his faded dreams that had died.

> *Damian Milton, Autistic scholar* (*1*)

### School Distress

School attendance problems driven by mental health (MH) difficulties are increasingly prevalent in the UK with the Children’s Commissioner’s recent Attendance Audit (“Where are England’s Children?” (2)) estimating that 1.7 million pupils in England were persistently absent (missing over 10% of school sessions) and 124,000 pupils severely absent (missing over 50% of school sessions) in the autumn 2021 term.

SAPs underpinned by MH difficulties are, however, not a new phenomenon. Failure by the scientific community to agree a typology for describing SAPs driven by MH difficulties (3) has hindered understanding and support, and led to a phenomena that is poorly described in the literature (4). Terms such as “school refusal”, “school phobia” and “school avoidance” have been used interchangeably throughout the literature to describe SAPs, with “school refusal behaviour” frequently used as an umbrella term covering anxiety-based school refusal and truancy (3).

Terms such as school “refusal” and “avoidance” are, however rejected by many individuals with lived experience of SAPs driven by MH difficulties, as they suggest the behaviour is under the control of the young person. Moreover, they do not convey any information regarding the emotional distress associated with school attendance experienced by these children and young people (CYP).

The term “School Anxiety” has become increasingly prevalent over recent years. This term, whilst addressing the above concerns, is narrow, focusing only on the anxiety component of the CYP’s experience. This places the focus on treating the CYP’s anxiety, as opposed to simultaneously addressing the drivers of this anxiety, for example a CYP’s sensory distress that is driven by being in a loud, busy, and unpredictable school environment.

We propose that SAPs underpinned by emotional distress are best described as “School Distress” (SD), given that emotional distress associated with school attendance is the core driving feature. This term does not focus solely on the anxiety component of the phenomena, which may be the outcome, rather than the driver, of the distress experienced by the CYP at school. Unlike terms such as “School Refusal”, SD is person-orientated and, as such, attempts to convey information to individuals surrounding CYP with respect to the child’s experience and presentation. We hope that this will intrinsically foster greater understanding and earlier recognition (particularly to early signs of distress), ultimately leading to more empathetic, appropriate support for these CYP.

### Presentation and prevalence of SD

SD is considered equally common amongst boys and girls, and no socioeconomic differences have been noted between children who do and do not experience these difficulties (5). The onset of SD may be sudden or gradual, with possible presentations including children pleading to miss school, displaying physical refusal in the morning, or expressing somatic complaints (6, 7).

Although SD likely accounts for a significant proportion of school absences, official figures are not available in the UK, with absences due to MH difficulties not being recorded differently to general absences. This prevents a full estimation of the scale of the problem. However, even if available, these figures would exclude CYP experiencing SD who still manage to attend school but who experience significant distress whilst there. Some authors estimate SAPs due to emotional distress affect around 1% of school-aged children (8), although others suggest higher estimates (e.g. (6, 9)), likely due to the different conceptualisations used (10).

### Underpinnings

Within the current literature, limited research has directly explored the causes of SD. Despite this, some potential factors have emerged within small-scale interviews with CYP and their parents, including fear of teacher behaviour, noisy and disorganised classrooms, anxiety, isolation, and unpredictability (11–13). In recent years, autism and sensory processing difficulties have become increasingly recognised as common characteristics amongst CYP experiencing SAPs (14).

### Autism

Autism has traditionally been characterised in the clinical and psychological literature as a deficit in social communication and interaction, the presence of restricted and repetitive patterns of behaviour, interests, or activities, and differences in sensory processing (15), with sensory processing differences believed to occur in as many as 90% of autistic individuals (16, 17). However, contemporary research and autistic scholars argue that a paradigm shift away from this deficit-based model, and a more nuanced understanding, is needed; one that situates autistic communication differences within a wider social context and considers communication breakdowns to be a consequence of differing perspectives between autistic and non-autistic individuals (18, 19).

Strikingly, Ochi et al. (20) found 40% of their school ’refusing’ participants to be autistic. Given that the prevalence of autism is 1-2% (21), such figures highlight an increased risk of SAPs (and likely also SD) in autistic children. Moreover, Munkhaugen et al. (22) identified teacher-reported SAPs in 42.6% of autistic students, compared to 7.1% of neurotypical (NT) students. Notably, this difference persisted when primary and secondary students were studied separately, indicating autistic students are at heightened risk of SAPs across their school life. Furthermore, autistic CYP displayed SAPs on a significantly greater number of days than their neurotypical counterparts, indicating greater severity, aligning with Ochi et al. (20) who reported a significantly lower mean age of onset of SAPs in autistic than nonautistic children.

Hence, there may be something specific about autistic CYPs’ experience of school that increases risk of experiencing SAPs and the severity of these difficulties, thereby attenuating the age at which difficulties begin and increasing duration. If this is the case, and particular groups of individuals with a recognised disability are specifically affected by SD, then this is of grave concern, not least when one considers the Equality Act (23), Children Act (24), Articles 23 and 28 of the United Nations Convention on the Rights of a Child (25), and Protocol 1, Article 2 of the Human Rights Act (“No person shall be denied a right to an education”) (26).

Insights into why autistic CYP have disproportionately negative experiences at school, and thus why they may be at increased risk of SD, are available from multiple sources (27–32). Contributing factors overlap with previously identified drivers (11–13) and include sensory processing difficulties, feelings of exclusion, lack of teacher understanding, and anxiety.

### Special Educational Need

Importantly, several of the above contributing factors are not specific to autistic CYP. For instance, D’Alessio (33) highlighted that CYP with Special Educational Needs (SEN) more broadly are frequently segregated from peers, resulting in feelings of exclusion within mainstream environments. This is supported by more recent work by Webster et al. (34) who described the experience of high-needs SEN CYP as often equating to “marginalisation masquerading as mainstream” (p. 77). Thus, it is possible that high-needs SEN CYP more generally will be over-represented amongst CYP experiencing SD due to the exclusion and marginalisation faced when attending mainstream schools, and the negative impact this may have on their wellbeing.

### Sensory Processing Differences

Sensory processing difficulties likely also play a contributing role (28). Sensory processing difficulties can affect one or multiple sensory systems, with these systems being hyper-reactive, hypo-reactive, or alternating between both states (35). There is increasing evidence that sensory processing difficulties affect CYPs’ school experiences, with mainstream school environments often consisting of “sensory exclusion” that disadvantage and marginalise autistic CYP (36). In support of this, parents in Havik et al.’s study (12) highlighted noisy classrooms as a contributing factor to SD, and McDougal et al. (32) reported that teachers with experience teaching autistic children in mainstream and SEN provisions identified sensory issues as a key barrier to learning in classroom settings. Furthermore, Jones et al. (28) found that negative sensory experiences in school can impact learning, cause distraction and anxiety, and limit participation in education.

As described above, sensory processing differences are an area of important difference between autistic and non-autistic individuals’ lived experience (37), and many autistic CYP experience significant sensory processing difficulties (38). This could explain the increased prevalence of SD amongst autistic CYP. However, non-autistic neurodivergent CYP (such as CYP with ADHD) also experience sensory processing differences (39), as do many other CYP, such as CYP born prematurely (40). Hence both autistic and non-autistic CYP who experience SPD may be at heightened risk of experiencing SD.

### Anxiety

In addition, and common across empirical studies, is the observation that anxiety plays an important role in the emergence and/or persistence of SD for many CYP [e.g., (13)], with high anxiety levels commonly noted in CYP experiencing SAPs [e.g., (41)]. For example, Gonzalvez et al. (42) used the DASS-21 to explore anxiety levels in three groups of CYP: CYP exhibiting ’school refusal behaviour’ (SRB) to avoid negative affectivity, escape social and/or evaluative situations, or pursue attention from significant others, CYP exhibiting SRB to pursue tangible reinforcements outside of school, and CYP exhibiting no SRB. Notably, the former group had significantly higher anxiety than CYP exhibiting no SRB and CYP who engaged in SRB to pursue tangible reinforcements. This is relevant here as the SRB exhibited by the former group appears to be most in line with the phenomenon we have described as SD, such that the behaviour is driven by the avoidance of aversive/distressing situations in school. Moreover, Jones et al., (43) found significantly greater clinician- and child-reported anxiety severity amongst school-reluctant CYP, compared to non-school reluctant CYP. Thus, high anxiety appears to be another characteristic of CYP experiencing SD. Of note, however, these studies do not tell us whether high anxiety is a cause or a consequence of CYPs’ distressing experiences in school.

Notably, severe symptoms of anxiety frequently co-occur in autistic individuals (44). Hence, understanding the role of both autism and anxiety, and the intersection between the two, in the development and maintenance of SD is likely important in elucidating key drivers of SD in CYP. Similarly, other ND conditions that also frequently co-occur with anxiety may also heighten the risk of experiencing SD [e.g., ADHD (45)], as, theoretically, could a primary diagnosis of anxiety.

### Demand Avoidance

Notably, parents of CYP experiencing SAPs often highlight their child’s difficulties coping with everyday demands as being instrumental in the difficulties faced at school, leading to extreme distress and/or behaviours (46). Such demands are omnipresent in the adult-directed mainstream setting, indicating a potential link between SD and demand avoidance.

Pathological Demand Avoidance (PDA) was first described by Newson as a distinct subtype within the diagnostic category of Pervasive Developmental Disorders (46, 47), and described CYP who displayed seemingly “obsessive resistance” to everyday demands and an extreme need for control (48). More recently, it has been suggested PDA may be more appropriately re-termed as ‘Extreme Demand Avoidance’ (EDA) (49), or should be fundamentally re-conceptualised as ’Rational Demand Avoidance’ (RDA) [i.e., as a rational response to one’s circumstances (1, 50)].

Although research is limited, a population cohort study from the Faroe Islands suggested almost 1 in 5 autistic CYP may show some demand avoidant characteristics (49). However, despite Newson’s work in the 1980s, PDA does not currently appear as a clinical diagnosis in the 5th edition of the Diagnostic and Statistical Manual of Mental Disorders (DSM-5) (15) or in the International Classification of Diseases 11th Revision (51). However, it is recognised by the National Autistic Society (NAS) in the UK as a variant of autism (52), and “demand avoidant behaviours” are referenced in the National Institute for Health and Care Excellence (NICE) clinical guidance for autism recognition, referral and diagnosis (53). Recent research has indicated that extreme demand avoidant behaviours in adults may be anxiety-driven (54), and that demand avoidant behaviour in CYP may be, in part, an attempt to increase certainty and predictability in order to alleviate increasing anxiety (55).

With respect to SD, a key motivation for Newson’s recognition of PDA in the 1980s was that a lack of recognition of this “markedly divergent overall presentation…contributes to inappropriate handling and educational methods, since PDA children respond best to very different approaches compared with those suitable for autistic and Asperger children”. More recently, Summerhill and Collett (56) highlighted anecdotal evidence indicating that when demand avoidant CYP are not identified in a timely manner, their presentation is viewed by others as defiance and deliberately challenging behaviour, leading to school exclusions (57). This, in turn, negatively impacts these CYP’s access to education, social relationships, and MH (56).

The uniqueness of the demand avoidant profile, coupled with its links to anxiety, may explain concerning recent findings for demand avoidant autistic CYP with respect to access to school-based education. For instance, a 2018 survey found that 70% of school aged demand avoidant CYP were either not enrolled in school or were unable to tolerate their school environment (58). Additionally, Truman et al. (59) found that whilst all parents of autistic CYP described their child’s school experiences as overwhelmingly negative, parents of demand avoidant autistic CYP provided markedly more negative descriptions than parents of autistic CYP without demand avoidant profiles. Hence, demand avoidant autistic children may be especially vulnerable to SD, perhaps due to their elevated levels of anxiety (60) and need for alternative educational methods (46, 47), which likely require a flexible, non-directive teaching style (61).

Interestingly, demand avoidance is not a profile exclusive to autism, but has also been documented in other neurodivergent profiles such as selective mutism, language disorders, epilepsy, and, less commonly, in the general population (49). Thus, demand avoidance may also play a role in SD in non-autistic neurodivergent and neurotypical CYP.

Understanding how demand avoidance relates to SD, and the parameters discussed above (e.g., anxiety, sensory processing differences), is thus important in order to fully elucidate the factors that contribute to SD. To date, however, there is a notable dearth of academic research exploring the link between demand avoidant profiles and SD. Understanding this is also relevant to informing best practice in supporting demand avoidant CYP to access education, as pressure to comply with direct demands is well-documented to lead to escalation in emotional reactivity and challenging behaviour in demand avoidant CYP (62).

### Aims and Hypotheses

By comparing CYP who have experienced SD with CYP who attend school without distress, and with CYP who have never attended a school setting [i.e., Electively Home-Educated (EHE) CYP], we aimed to:

1. **Quantify the proportion of cases of SAPs which are associated with emotional distress.**
2. **Identify prevalent characteristics of CYP who have experienced difficulties attending school, including the prevalence of a wide range of common neurodivergent profiles, and mental and physical health difficulties.**
3. **Compare anxiety levels, sensory processing difficulties, and demand avoidance profiles in CYP who experience SD and in those who do not.**
4. Explore associations between sensory processing difficulties, anxiety, demand avoidance, and markers of SD severity (i.e., duration of SD, school attendance rate, age of SD onset, and impact of school attendance on MH).
5. **Explore the number and types of education settings attended by CYP with SD.**
6. **Assess the level of support received by CYP currently experiencing SD.**

It is hypothesised that neurodivergent (ND) CYP will be overrepresented amongst individuals with SD experience, particularly autistic CYP and CYP with sensory processing difficulties; that anxiety will be prevalent in CYP with SD, particularly in autistic, non-autistic ND, and/or demand avoidant CYP; and CYP with more extensive sensory processing difficulties, higher anxiety, and more pervasive demand avoidant profiles, will show more severe SD than their neurotypical peers. Additional research questions will be addressed elsewhere.

## Materials and Methods

### Participants

Participants were required to live in the UK and be parents/carers of school-aged CYP. Initially, 1055 participants were recruited via volunteer sampling, consisting of 738 parents of children currently experiencing SD (Current SD), 209 parents of children who have previously experienced SD (Past SD), 83 parents of children who have never experienced SD (No SD), and 25 parents of children who have never attended a school setting for reasons other than SD (Lifelong EHE). An additional 66 parents of CYP who have never experienced SD were recruited via prolific.org to ensure the Current, Past, and No SD groups were all matched in terms of chronological age, providing an overall sample of 1121 participants. To assist with age matching, prolific parents with more than one child were instructed to consider their eldest child within the questionnaire. On average, participants completed 77.35% of the survey, with 62.5% completing 100%. Most participants were mothers (97.03%). Table 1 displays key characteristics of the CYP. Figure 1 shows a map of the CYP experiencing SD, by county.

**Table 1.** Demographic characteristics of the sample (N=1121)

| Characteristic | All | Current SD | Past SD | No SD | Lifelong EHE |
| --- | --- | --- | --- | --- | --- |
| *n* (%) | 1121 | 738 (65.8) | 209 (18.6) | 149 (13.3) | 25 (2.2) |
| Respondent's Relationship to Child (%) |  |  |  |  |  |
| Mother | 1077 (97.0) | 707 (96.6) | 200 (98.0) | 145 (97.3) | 25 (100) |
| Father | 18 (1.6) | 14 (1.9) | 1 (0.5) | 3 (2.0) | 0 |
| Other | 15 (1.4) | 11 (1.5) | 3 (1.5) | 1 (0.7) | 0 |
| Mean Age in Years ± SD | 11.6 ± 3.3 | 11.8 ± 3.1 | 11.8 ± 3.6 | 11.1 ± 3.5 | 8.7 ± 3.0 |
| Gender (%) |  |  |  |  |  |
| Cisgender Boy | 577 (52.1) | 381 (52.1) | 117 (57.6) | 68 (45.6) | 11 (44.0) |
| Cisgender Girl | 471 (42.5) | 300 (41.0) | 77 (37.9) | 80 (53.7) | 14 (56.0) |
| Transgender Boy | 9 (0.8) | 9 (1.2) | 0 | 0 | 0 |
| Transgender Girl | 1 (0.1) | 0 | 1 (0.5) | 0 | 0 |
| Non-binary | 27 (2.4) | 23 (3.1) | 4 (2.0) | 0 | 0 |
| Self-describe | 11 (1.0) | 8 (1.1) | 3 (1.5) | 0 | 0 |
| Prefer not to say | 12 (1.1) | 10 (1.4) | 1 (0.5) | 1 (0.7) | 0 |
| IMD Decile | 6.04 (2.9) | 6.16 (2.8) | 5.51 (3.1) | 6.17 (2.9) | 5.50 (2.6) |
| Ethnic Group (%) |  |  |  |  |  |
| White | 693 (93.4) | 458 (93.5) | 113 (95.8) | 106 (90.6) | 16 (94.1) |
| Mixed/Multiple Ethnic Groups | 37 (5.0) | 24 (4.9) | 4 (3.4) | 9 (7.7) | 0 |
| Asian/Asian British | 6 (0.8) | 3 (0.6) | 1 (0.8) | 1 (0.9) | 1 (5.9) |
| Black/African/Caribbean/Black British | 3 (0.4) | 2 (0.4) | 0 | 1 (0.9) | 0 |
| Other Ethnic Group | 3 (0.4) | 3 (0.6) | 0 | 0 | 0 |
| Main Language (%) |  |  |  |  |  |
| English | 735 (99.3) | 484 (99.2) | 117 (100) | 117 (99.2) | 17 (100) |
| Other | 5 (0.7) | 4 (0.8) | 0 | 1 (0.8) | 0 |
| Country of Residence (%) |  |  |  |  |  |
| England | 980 (88.3) | 644 (88.0) | 183 (89.7) | 129 (86.6) | 24 (96.0) |
| Scotland | 94 (8.5) | 68 (9.3) | 13 (6.4) | 13 (8.7) | 0 |
| Wales | 22 (2.0) | 14 (1.9) | 5 (2.5) | 2 (1.3) | 1 (4.0) |
| Northern Ireland | 14 (1.3) | 6 (0.8) | 3 (1.5) | 5 (3.4) | 0 |
| Siblings (%) |  |  |  |  |  |
| Yes | 850 (79.7) | 571 (81.1) | 145 (75.9) | 118 (79.2) | 16 (69.6) |
| No | 217 (20.3) | 133 (18.9) | 46 (24.1) | 31 (20.8) | 7 (30.4) |
| Position in Family (%) |  |  |  |  |  |
| Youngest | 329 (33.6) | 250 (39.2) | 54 (30.5) | 20 (14.1) | 5 (22.7) |
| Middle | 92 (9.4) | 68 (10.7) | 17 (9.6) | 5 (3.5) | 2 (9.1) |
| Eldest | 314 (32.1) | 169 (26.5) | 57 (32.2) | 81 (57.0) | 7 (31.8) |
| Twin | 27 (2.8) | 18 (2.8) | 3 (1.7) | 5 (3.5) | 1 (4.5) |
| Only Child | 217 (22.2) | 133 (20.8) | 46 (26.0) | 31 (21.8) | 7 (31.8) |
| Academic Attainment |  |  |  |  |  |
| Not Meeting Expectations | 543 (52.6) | 462 (67.6) | 65 (36.1) | 12 (8.1) | 4 (18.2) |
| Meeting Expectations | 323 (31.3) | 151 (22.1) | 74 (41.1) | 85 (57.4) | 13 (59.1) |
| Exceeding Expectations | 167 (16.2) | 70 (10.2) | 41 (22.8) | 51 (34.5) | 5 (22.7) |
| SEN Support (*\*Mainstream Only*) |  |  |  |  |  |
| No SEN Support* | - | 152 (35.9) | 36 (100) | 122 (97.6) | n/a |
| SEN Support* | - | 194 (45.5) | 16 (100) | 15 (10.6) | n/a |
| EHCP/Statement in place* | - | 91 (21.2) | 19 (26.4) | 6 (4.2) | n/a |
| EHCP/Statement in progress* | - | 64 (14.9) | 0 | 0 | n/a |

This study was approved by the Faculty of Medical Sciences Research Ethics Committee, part of Newcastle University’s Research Ethics Committee.

### Language

Where possible, we use identity-first language (e.g., autistic CYP) (63). We defined ND as “a term for when someone’s brain processes, learns, and behaves differently from what is considered ‘typical’. Autism is an example of a neurodivergence”. We use the term ‘non-autistic’ to refer to CYP whose parents did not identify them as autistic (be that diagnosed or self/parent-identified), and ‘non-autistic ND CYP’ for the subgroup of non-autistic CYP who are otherwise ND. We use the term ‘neurotypical’ (NT) to refer to CYP whose parent identified them as not being neurodivergent.

### Design

The study employed a case-control, concurrent embedded, mixed-methods design where qualitative data was collected to supplement quantitative data. This was chosen due to the study’s exploratory nature, and because the limited literature prevented us from providing fully comprehensive lists of response options to some questions. To collect qualitative data, text boxes were presented within some questions for parents to provide comments. The results reported in this paper are largely quantitative, with some parental comments reported to support understanding. Thematic analyses and additional data will be reported elsewhere.

### Materials

A bespoke online questionnaire was developed containing four sections and 76 questions. Only certain questions were presented to each respondent, based on their experience of SD and survey responses. Questions and response options were developed based upon a comprehensive literature review, and aimed to collate key information about the respondent, their CYP, their CYP’s experience of SD, and the impact of SD on themselves.

This paper will report data from the questions and clinical scales described below. Data relating to how SD presents, the reasons underlying SD, the efficacy of supports, the consequences of SD, and the parental experience will be reported elsewhere.

*1. Demographic Information*: Participants were asked their relationship to the CYP, and the CYP’s country of residence, spoken language, ethnicity, age, gender identity, and number of siblings (with ages). Postcodes were requested and converted into Index of Multiple Deprivation (IMD) Deciles for families living in England (64). In accordance with prolific.org data protection policy, prolific participants provided only their IMD decile (using https://www.fscbiodiversity.uk/imd/).
*2. School Attendance Problems*: Participants were asked whether their child has ever experienced difficulties attending school (response options: ‘currently’, ‘in the past’, ‘never’, or ‘not applicable as child never attended a school setting’), and if so, what age their difficulties began. Where attendance difficulties were current, parents were asked how long they had been ongoing, and how many days their CYP had attended school over the proceeding 20 school days. Attendance rates for the current (2021/22) and previous academic year (2020/21; excluding Covid-19-related absences) were also estimated. All parents of CYP with SAPs were asked to describe these difficulties using one of the following options: “Self-corrective school avoidance (i.e. absenteeism that remits spontaneously within 2 weeks)”; “Acute school avoidance (i.e. absenteeism that lasts from 2 weeks to 1 year)”; “Chronic school avoidance (i.e. absenteeism that lasts longer than 1 year)”; “None of the above. It looks more like…(please describe)”. In addition, all parents were asked about the impact of attending school on their child’s MH [response options: ‘Extremely positively’ (+3), ‘Very positively’ (+2), ‘Somewhat positively’ (+1), ‘Neither positively nor negatively’ (0), ‘Somewhat negatively’ (-1), ‘Very negatively’ (-2), ‘Extremely negatively’ (-3)]. Detailed results will be presented elsewhere, however this measure will be used here as a marker of SD severity. Finally, parents were also asked whether the CYP’s siblings have a history of SAPs.
*3. Educational Information*: Parents were asked to indicate the types of educational provision their child currently (and if relevant, previously) attended, the total number of schools their child had attended, and whether their child was currently receiving SEN support at school [response options: ‘receives no additional support’, ‘receives SEN support (e.g. is on the SEN register)’, or ‘has an EHCP/Statement/CSP/ALN (or similar) in place or in process’].
*Child Health and Neurodivergencies*: To better understand the needs of CYP struggling to attend school, parents were asked if their child has any physical or MH difficulties (and, if so, what they were), and if they are ND. Parents who stated their child was (or might be) ND were provided with a list of possible neurodivergencies (i.e., Autism, ADHD, Auditory Processing Disorder (APD), Dyscalculia, Dyslexia, Dyspraxia, Gifted, Intellectual Disability, Language Disorder, Sensory Processing Disorder/Sensory Integration Disorder (SPD/SID), Speech Difficulties, Tic Disorder, Unspecified Learning Disorder, Visual Processing Difficulties, Other) and were asked to select all that apply to their child (response options: ‘has a clinical diagnosis’, ‘is on the diagnostic pathway’, ‘had a referral refused’, ‘suspected but has never been referred and/or diagnosed’). Unless otherwise indicated, prevalence rates for each ND were calculated by accepting endorsement of any of these four options. Parents who identified their CYP to have SPD/SID were presented with the eight sensory systems (Visual, Auditory, Tactile, Olfactory, Gustation, Vestibular, Proprioceptive, and Interoceptive) and asked to identify those in which their CYP experienced difficulties. Parents who selected Intellectual Disability were asked if this was best described as ‘mild-moderate’, ‘severe’, or ‘profound’. To gain a wider understanding of the CYP’s family history, we also asked whether either of the CYP’s parents, or their siblings, are ND. Rates of parental ND will be published elsewhere. All parents were asked to complete the 24-item Anxiety Scale for Children–Autism Spectrum Disorder–Parent Version (ASCASD-P) (65), which is derived from a well-validated measure used with typically-developing children (66) and developed for use with autistic CYP. The ASC-ASD-P was selected given the anticipated rates of autistic CYP in our sample. This parent-report measure provides a total anxiety score, and individual scores for Separation Anxiety, Uncertainty, Performance Anxiety, and Anxious Arousal (total anxiety will be presented here, and findings regarding specific subscales will be reported elsewhere). Parents respond along a 4-point Likert scale (0=never, 3=always), and scores are calculated by summing responses, with higher scores indicating greater levels of anxiety (range 0-72). A total score of 20-23 suggests “significant anxious symptomatology”, and scores above 24 suggest a “more specific indication of significant anxiety” (65). The ASC-ASD-P has excellent internal consistency (α=0.94) and good convergent validity.
*5. Demand Avoidance*: All parents were asked to complete the 8item Extreme Demand Avoidance-8 Caregiver Report Questionnaire (EDA-8) (67), which is a refined version of the Extreme Demand Avoidance Questionnaire (EDA-Q) (68). The scale’s items cover features consistently described in accounts of EDA: obsessive avoidance of demands and requests, outrageous or shocking behaviour to avoid, need for control, poor awareness of hierarchy, and lability of mood. The EDA-8 has good internal consistency (α=.90) and convergent and divergent validity, and is proposed to be a useful tool to identify children showing an extreme response to demands (67). Parents respond along a 4point Likert scale (0=Not at all true, 3=Very true). Scores are calculated by summing responses, with higher scores indicating greater EDA. Cut-off scores are not currently available.

### Procedure

Data was collected using Qualtrics. The survey link was shared widely on social media, and the additional control participants recruited via prolific.org were directed to the Qualtrics link.

Participants read the information sheet and provided consent before beginning the survey. They were informed they could skip any questions and stop/start at any time. Qualtrics’s display-logic function ensured respondents were only asked questions which were relevant to them. Upon completion, participants were presented with a debrief form, including a comprehensive list of support services. The study ran for 14 days (22/02/2022-08/03/2022).

### Data Analysis

Participants were designated to one of four groups based upon whether their child had ever experienced difficulties attending school: the response option ’Yes, currently’ assigned them to the Current SD group, ’Not currently, but they have in the past’ to the Past SD group, ’No, never’ to the No SD group, and ’Not applicable as child never attended a school setting’ to the Lifelong EHE group.

Quantitative data analyses were run using IBM SPSS Statistics V26. Descriptive statistics were calculated to summarise participants’ responses to each question. Further statistical analyses were conducted to examine relationships between variables. Before performing statistical analyses, normality was assessed by plotting results in histograms and conducting Shapiro-Wilk and Kolmogorov-Smirnov tests. When results were not normally distributed, non-parametric methods were used. A significance level of **α**=0.05 was adopted for most analyses, and a Bonferroni-corrected alpha level was used to correct for multiple comparisons. Odds Ratios (ORs) were calculated as an estimate of effect size.

As CYP in the Lifelong EHE group were significantly younger than CYP in the other groups [(Current SD=Past SD=No SD)>Elective EHE, p<.001], it was necessary to conduct additional analyses using more precisely age-matched comparison groups. Hence, for each young person in the Lifelong EHE group, two aged-matched participants were identified from each of the three other groups. The selected CYP from each group were the two CYP closest in age to the corresponding Lifelong EHE young person. Analyses were then replicated using this reduced sample, and conclusions specific to the Lifelong EHE group were derived from these results.

Within our study, we measured four key proxy markers of SD: SD Duration, age of onset of SD, attendance rates (in the previous 20 school days, 21/22 academic year, 20/21 academic year), and impact of school attendance on MH. Within this paper, we explored relationships between the four proxy markers of SD and several characteristics of our sample (i.e., anxiety, DA, and the number of sensory systems that CYP experience difficulties in) using Spearman rho correlations to see how they related to SD severity.

## Results

### 1. Demographic Information

For summary of demographic information, see Table 1.

#### Gender

52.1% of the CYP in the sample were identified by their parents as cisgender boys, 42.5% as cisgender girls, 2.4% as non-binary, 0.8% as transgender boys, 0.1% as transgender girls, and the remaining 2.1% were split between the “self-describe” and “prefer not to say” options.

#### Current Age

The mean age of the CYP in our sample was 11.6 years. As described above, there was an overall between-group difference with respect to current age [F(3, 1106)=8.548, *p*<0.001]. Post-hoc analyses indicated that this difference was driven by CYP in the Lifelong EHE group who were significantly younger than the three other groups [Current SD versus Past SD: *p*=1.0; Current SD versus No SD Control: *p*=0.159; Past SD versus No SD Control: *p*=0.348; Lifelong EHE versus No SD Control: *p*=0.004]. No differences remained for the Lifelong EHE age-matched subgroup ([F(3, 171)=0.084, *p* = .969].

#### Indices of Deprivation

In total, Index of Multiple Deprivation Decile data was available for 348 families (47%) in the Current SD group, 97 families in the Past SD group (53%), 88 families (59%) in the No SD group, and 4 families (16%) in the Lifelong EHE group. Based on this data, there were no significant between-group differences with respect to the Index of Multiple Deprivation (IMD) Decile scores [F(3, 533)=1.413; *p*=.238].

A one sample t-test compared the mean IMD Decile against the population mean, estimated to be 5.5. The overall mean in our sample was 6.04 (StDev=2.9), which was significantly higher than the population mean, t(536)=4.312, p<.001, indicating less deprivation. Broken down into groups, the Current (6.16, StDev=2.82) and No SD group means (6.17, StDev=2.96) were significantly higher than the population mean [Current SD: t(347)=4.368, p<.001, No SD: t(88)=2.127, p<.001], whilst the Past SD (5.50, StDev=3.09) and Lifelong EHE group means (5.50, StDev=2.65) were not significantly different than the population mean [Past SD: t(96)=0.16, p=.987, Lifelong EHE: t(3)=0.000, p=1.000].

#### Birth Order

47.3% of Current SD CYP were first-born children (26.5% ‘eldest’ children and 20.8% an ‘only’ child), relative to 49.9% who were younger siblings (39.2% ‘youngest’ and 10.7% ‘middle’ children). Notably, when considering CYP who had experience of SD and who were either the ‘youngest’ or a ‘middle’ child in their family, we found having an older sibling who had also experienced SD was common. More specifically, 42.9% of younger siblings in the Current SD group, and 46.5% of younger siblings in the Past SD group, also had an older sibling/s with a history of SD. Similar figures were obtained when ‘youngest’ children were considered in isolation (see Figure S1).

### 2. School Attendance Difficulties

#### Age of Onset and Duration of SAPs

The mean age of onset of SD across both groups was 7.89 years (StDev=3.37). This was younger in Past SD CYP (7.19 years; StDev=3.21) than Current SD CYP (8.07 years; StDev=3.21). Strikingly, 51.2% of cases of SD first occurred at 8 years or younger. The mean duration of SD was 3.99 years (StDev=2.95). This was longer for CYP whose difficulties had now resolved (4.79 years; StDev=3.12), than for those whose difficulties were still ongoing (3.79 years; StDev=2.88). Hence, SD began significantly earlier in Past SD CYP (p<.01), and lasted significantly longer (p<.001), likely because the Current SD CYP were still experiencing SD. Notably, the age of onset of SD was significantly younger for autistic CYP than non-autistic CYP, and SD was reported as being significantly more enduring for autistic CYP (see Figure 2).

**Fig. 1.**
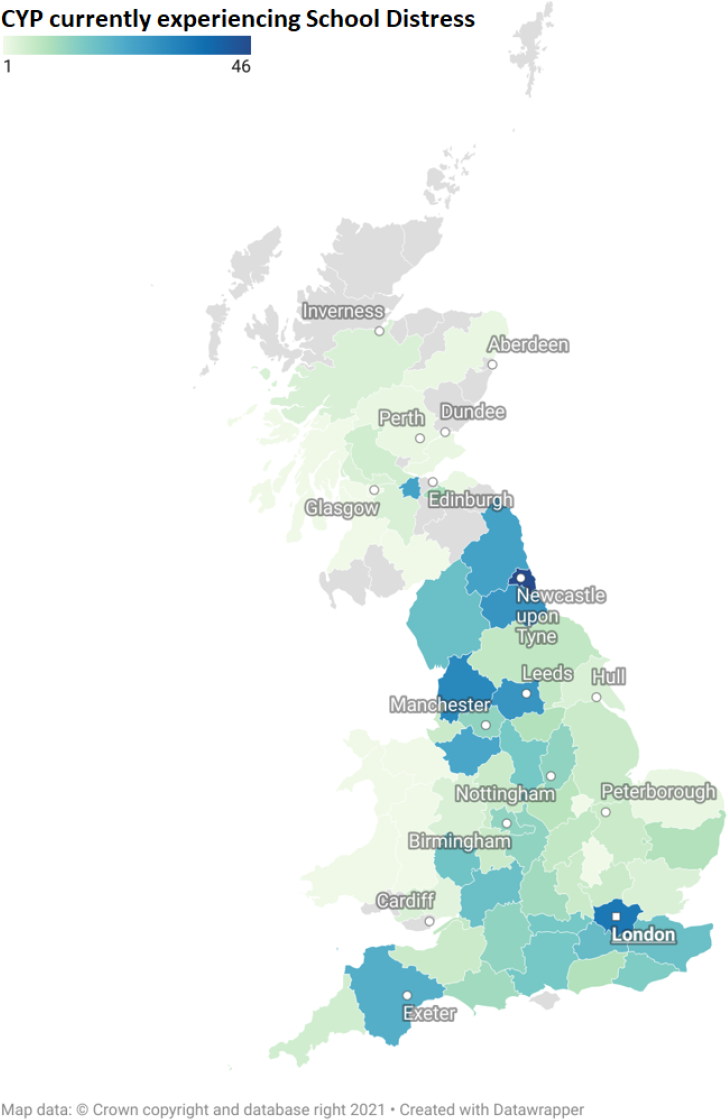
Map of CYP currently experiencing School Distress, excluding Northern Ireland

**Fig. 2.**
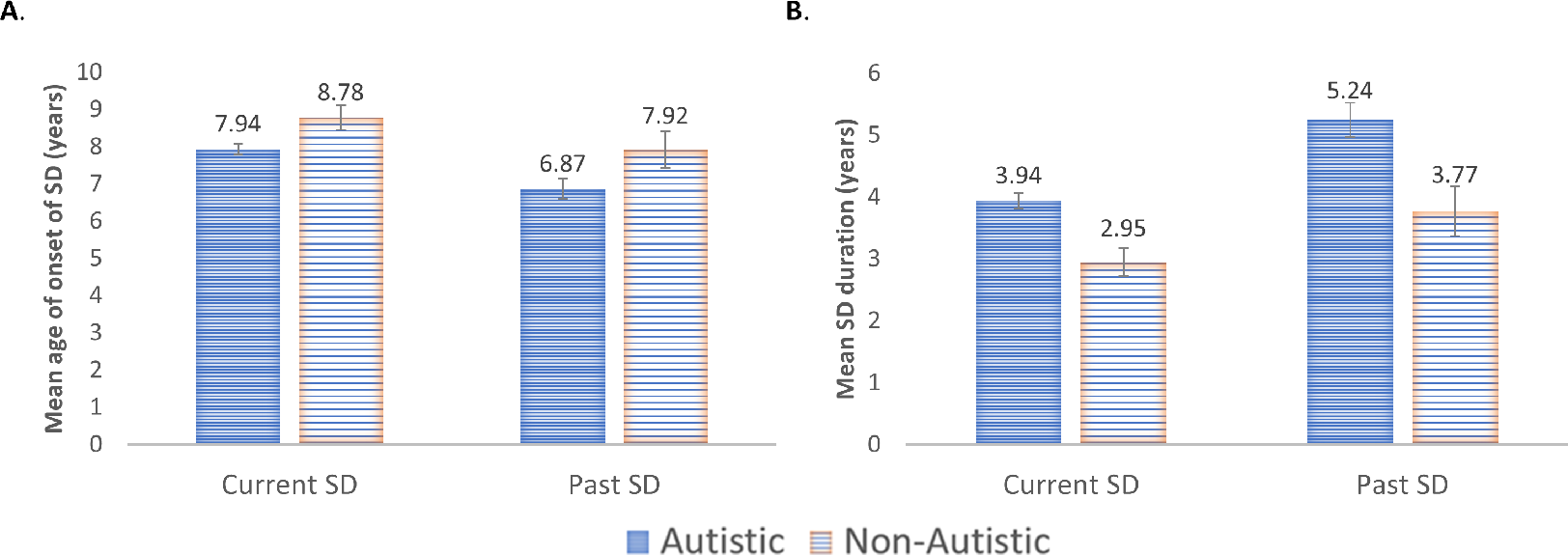
Panel A: Age of Onset of SD. Panel B: Duration of SD. Error bars: ±1 SEM

Given the timing of this research, with survey completion occurring almost 2 years after the initial Covid-19 school closures began in the UK, we explored the percentage of SD cases that began before and after the Covid-19 pandemic. In most cases, the onset of SD preceded Covid-19 related school closures (Current SD: 69.82%; Past SD: 85.15%).

#### School Attendance Problems

For 94.32% of CYP in the sample, parents indicated that their child’s SAPs were either partially or fully emotionally based (i.e., SD).

The existing categories for school “refusal” within the literature (58) (self-corrective, acute, and chronic) failed to capture a significant proportion of the experiences of CYP in this sample (see Table 2, Other), with 37.1% of cases (n=320) falling outside of these categories. Examining the Past SD group alone, the ‘none of the above’ category was selected by 54.2% of parents. Parents who selected this option were asked to describe what their child’s SD looks like. Example responses can be seen in Table 3, Q1. These descriptions capture the distress element of the lived experience of SD, as do the additional quotes with respect to how SD presents in their children (Table 3, Q2). A full thematic analysis of this data will be described elsewhere.

**Table 2.** Type of School Attendance Problems.

| Variable | Combined SD | Current SD | Past SD |
| --- | --- | --- | --- |
| Type of School Attendance Problems (%) |  |  |  |
| Self-corrective | 134 (15.55) | 105 (15.37) | 29 (16.20) |
| Acute | 198 (22.97) | 172 (25.18) | 26 (14.53) |
| Chronic | 210 (24.36) | 183 (26.79) | 27 (15.08) |
| Other | 320 (37.12) | 223 (32.65) | 97 (54.19) |
| Emotionally Based School Attendance Problems (%)<sup>a</sup> | 813 (94.32) | 648 (94.88) | 165 (92.18) |

<sup>a</sup>Reflects the number and percentage of parents who reported that their child's school attendance problems either fully or partially emotionally based.

**Table 3.**
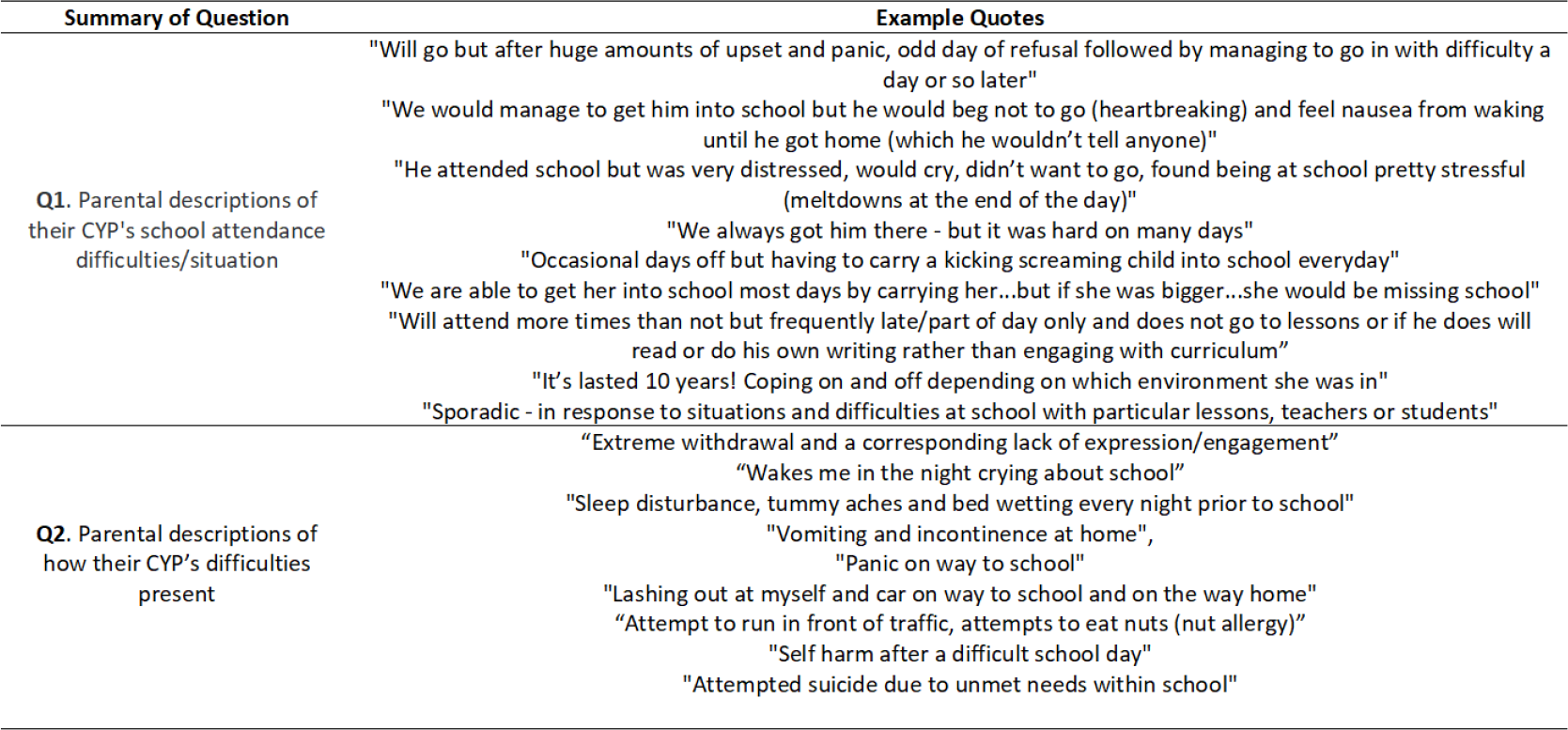
A sample of quotations provided by parents in response to specific questions.

### 3. Educational Information

#### Type of Education Setting Attended

Overall, 97% of CYP had previously, or were currently, attending a mainstream school setting (Current SD: 97%, Past SD: 97%, No SD: 99%). Almost all CYP in the No SD group were currently attending a mainstream school, whilst just 58.3% of CYP in the Current SD group remained in this setting currently. The current and past educational provisions of CYP with SD experience will be described in more detail elsewhere.

The average number of schools attended by CYP was 2.24 (StDev=1.085, range 1-6) [Current SD=2.36 (StDev=1.094), Past SD=2.22 (StDev=1.164), No SD=1.86 (StDev=0.814)].

Notably, there was a significant between-group difference [F(2,1007)=12.986, p<.001], with the No SD group attending significantly fewer schools than the SD groups (Current SD>No SD, p<.001; Past SD>No SD, p=.009; Current SD=Past SD, p=0.360).

#### Support at School (SEN/EHCP)

Of the Current SD CYP, 32.8% received no support at school, 38.1% were on a SEN register (or equivalent), and 48.5% had an EHCP or were in the process of seeking one. This declined to 32.9% when we removed cases where parental comments indicated an EHCP was not yet in place (n=111). Of those cases, 95 parents indicated they were in the EHCP process [e.g., applying, applied, in the assessment phase, in the draft stage/awaiting a finalised plan, at mediation/appealing…]. Some of the mediations/appeals were taking place following a refusal to assess or to issue an EHCP following assessment, whilst others were appealing the content of sections B (description of the CYP’s SEN), F (provision required to meet needs), and I (specific placement).

Parental dissatisfaction with the support their child is/was receiving from their school and/or LA was clear throughout responses. For instance, many described a lack of support in place (e.g., “Very limited support from school”; see Table S1, Q1). Even when parents indicated their child was on the school’s SEN register or had an EHCP, comments continued to indicate a lack of support for many CYP (e.g., “Is on the SEN register however no further support in school”, see Table S1, Q2). Application for and implementation of EHCPs was also a particular source of frustration, with comments including: “I’d to self-apply as school delayed and blocked” and “School not following EHCP” (see Table S1, Q3).

Occasionally comments reflected a more positive situation (e.g., “My child’s school currently provides reasonable adjustments for my daughter’s needs “, see Table S1, Q4), however this only represented a small proportion of parent voices. Finally, some parental comments reflected the complexity of providing support (e.g., “He can receive support from a learning base but he is masking in school and doesn’t want others to know he has autism so only accesses the base twice per week for 50 mins per time”).

### 4. Child Health and Neurodivergences

#### Child Health

When asked whether their child has any physical or MH difficulties, only 7% of parents in the Current SD group responded ‘No’, compared to 69.8% who stated ‘Yes’ and 23.3% who stated ‘Maybe’. This differed to the other groups, whereby 23.5% of Past SD parents, 79.9% of No SD parents, and 56% of Lifelong EHE parents responded ‘No’. When asked to specify details of these health difficulties, some parents listed neurodevelopmental conditions. As we later gathered detailed information about neurodivergent conditions, we excluded such responses from this analysis (e.g., autism, ADHD, PDA, and sensory processing differences).

Table 4 summarises the remaining responses with respect to the CYPs’ mental and physical health difficulties, with anxiety being the most frequently mentioned condition in all groups. Depression, Hypermobility, PTSD/trauma, and Low mood/emotional regulation difficulties were the next most common in the two SD groups, with few incidences in the No SD and Lifelong EHE groups. Hence, MH as opposed to physical health concerns (except for hypermobility) were the most frequently mentioned health difficulties by parents of children with SD experience. Notably, this cannot be considered an exhaustive list, as some parents did not complete this, and others may not have listed all health concerns.

**Table 4.** Frequency and Percentage (%) of Children Within Each Group Who Have A Range of Physical and Mental Health Difficulties, As Listed by Parents (as this was an optional free-text question, this should not be considered an exhaustive list).

| Physical/Mental Health Difficulty | Current SD (n=731) | Past SD (n=204) | No SD (n=149) | Lifelong EHE (n=25) |
| --- | --- | --- | --- | --- |
| Anxiety | 339 (46.37%) | 63 (30.88%) | 15 (10.07%) | 5 (20%) |
| Depression | 70 (9.58%) | 9 (4.41%) | 2 (1.34%) | 0 |
| Hypermobility/ Ehlers Danlos Syndrome | 46 (6.29%) | 14 (6.86%) | 1 (0.67%) | 0 |
| PTSD/Trauma | 37 (5.06%) | 9 (4.41%) | 0 | 0 |
| Low Mood/Emotion Regulation Difficulties | 37 (5.06%) | 2 (0.98%) | 1 (0.67%) | 0 |
| Obsessive Compulsive Disorder (OCD) | 24 (3.28%) | 6 (2.94%) | 1 (0.67%) | 0 |
| Low Self Esteem | 14 (1.92%) | 0 | 0 | 0 |
| Asthma | 12 (1.64%) | 2 (0.98%) | 1 (0.67%) | 0 |
| Genetic/Chromosomal Disorder (e.g., Down Syndrome, Fragile X Syndrome) | 12 (1.64%) | 2 (0.98%) | 0 | 0 |
| Eating Disorder / Difficulties / ARFID | 12 (1.64%) | 1 (0.49%) | 0 | 0 |
| Self-Harm | 12 (1.64%) | 0 | 0 | 0 |
| Attachment Issues/Disorder | 10 (1.37%) | 3 (1.47%) | 0 | 0 |
| Co-ordination Difficulties (including DCD) | 10 (1.37%) | 1 (0.49%) | 0 | 1 (4%) |
| Phobias (e.g., Agoraphobia, Emetophobia) | 10 (1.37%) | 0 | 0 | 1 (4%) |
| Unspecified Mental Health Difficulties | 10 (1.37%) | 0 | 0 | 0 |
| Selective Mutism | 9 (1.23%) | 2 (0.98%) | 0 | 2 (8%) |
| Suicidal Ideation/Suicide Attempts | 9 (1.23%) | 1 (0.49%) | 0 | 0 |
| Hearing Difficulties | 6 (0.82%) | 0 | 0 | 0 |
| Visual Impairment | 5 (0.68%) | 1 (0.49%) | 2 (1.34%) | 0 |
| Fine and/or Gross Motor Difficulties/Delays | 5 (0.68%) | 1 (0.49%) | 0 | 0 |
| Bladder/Bowel Difficulties (e.g., IBS) | 5 (0.68%) | 2 (0.98%) | 2 (1.34%) | 0 |
| Allergies | 5 (0.68%) | 1 (0.49%) | 2 (1.34%) | 0 |
| Sleep Difficulties/Disorder | 5 (0.68%) | 1 (0.49%) | 0 | 0 |
| Epilepsy | 4 (0.55%) | 1 (0.49%) | 0 | 0 |
| Anger Issues | 4 (0.55%) | 0 | 1 (0.67%) | 0 |
| Acid Reflux | 3 (0.41%) | 1 (0.49%) | 0 | 0 |
| Eczema | 3 (0.41%) | 0 | 1 (0.67%) | 0 |

Note: This table does not include responses including ND conditions and sensory processing difficulties as these were quantified more precisely in later questions. AFRID = Avoidant Restrictive Food Intake Disorder, DCD = Developmental co-ordination disorder, IBS = Irritable Bowel Syndrome, OCD = Obsessive Compulsive Disorder, PTSD = Post-Traumatic Stress Disorder.

Table 5 further subdivided these responses into three categories: CYP whose parents listed MH difficulties only, whose parents listed physical health difficulties only, and whose parents listed both mental and physical health difficulties. Rates of CYP whose parents reported physical health conditions in isolation were relatively low in all groups (see Table 5). However, having either MH difficulties in isolation, or in combination with physical health difficulties, was strikingly more common in both SD groups than in both the No SD and Lifelong EHE groups. No formal statistical analyses were conducted here as more precise data (including anxiety data gathered using a clinical scale) is described below. Co-occurrence between neurodivergent conditions and health difficulties is also discussed further below.

**Table 5.** Percentage of Physical and Mental Health Difficulties per Group.

|  | Physical Health | Mental Health | Both | None Listed |
| --- | --- | --- | --- | --- |
| Current SD | 3.73% | 67.01% | 14.48% | 16.27% |
| Past SD | 7.53% | 49.46% | 6.45% | 36.56% |
| No SD | 4.05% | 10.14% | 1.35% | 84.46% |
| Lifelong EHE | 8.33% | 25% | 0% | 66.67% |

Note: Neurodivergencies such as autism, ADHD, dyslexia and sensory processing difficulties were excluded as they were assessed separately. The "None Listed" category includes CYP who do not have any health difficulties AND CYP whose parents opted not to specifically describe these difficulties.

#### Neurodivergence

Most Current SD CYP were rated as ND by their parents (92.05%), compared to just 22.2% of those without SD experience (see Table 6; note: frequencies reflect the number of parents who responded “yes” or “maybe” to their child being ND). Combining “yes” and “maybe” responses into one category, a Chi-Square test revealed a significant difference in the frequency of ND CYP across the four groups, **χ**²(3,1098)=394.5, p<.001. Post-hoc analyses indicated Current SD CYP were significantly more likely to be ND than Past SD CYP, and Current and Past SD CYP were significantly more likely to be ND than No SD CYP (Current SD>Past SD>No SD). Notably, the OR for a CYP to experience SD if ND was 32.57 (95% CI 20.903, 50.762). Restricting the criteria of ND to just the CYP whose parents responded “yes” increased this OR to 42.25 (95% CI [24.53, 72.78]). Hence, ND CYP were significantly over-represented amongst CYP who experience SD.

**Table 6.** Frequency (%) of Individual Neurodivergencies in Each Group, and Average Number of Neurodivergencies per Group.

| Diagnosis | Current SD | Past SD | No SD | Lifelong EHE |
| --- | --- | --- | --- | --- |
| Neurodivergent | 666 (92.1) | 168 (83.6) | 33 (22.2) | 22 (88.0) |
| Autism | 598 (83.4) | 133 (66.2) | 25 (16.8) | 13 (52.0) |
| Sensory Processing Disorder/Sensory Integration Disorder (SPD/SID) | 406 (56.9) | 87 (43.3) | 10 (6.7) | 13 (52.0) |
| Attention Deficit Hyperactivity Disorder (ADHD) | 395 (55.3) | 87 (43.3) | 13 (8.7) | 12 (48.0) |
| Dyslexia | 181 (25.4) | 47 (23.4) | 8 (5.4) | 2 (8.0) |
| Dyspraxia | 177 (24.7) | 55 (27.4) | 12 (8.1) | 2 (8.0) |
| Auditory Processing Disorder (APD) | 130 (18.2) | 26 (12.9) | 5 (3.4) | 3 (12.0) |
| Speech Difficulties | 115 (16.1) | 21 (10.4) | 7 (4.7) | 4 (16.0) |
| Gifted | 103 (14.4) | 29 (14.4) | 6 (4.0) | 6 (24.0) |
| Other | 89 (12.5) | 16 (8.0) | 3 (2.0) | 3 (12.0) |
| Dyscalculia | 88 (12.3) | 19 (9.5) | 4 (2.7) | 0 |
| Language Disorder | 82 (11.5) | 20 (10.0) | 4 (2.7) | 2 (8.0) |
| Visual Processing Difficulties | 69 (9.7) | 19 (9.5) | 0 | 0 |
| Tic Disorder | 61 (8.5) | 13 (6.5) | 5 (3.4) | 1 (4.0) |
| Unspecified Learning Disorder | 60 (8.4) | 10 (5.0) | 1 (0.7) | 1 (4.0) |
| Intellectual Disability | 48 (6.7) | 10 (5.0) | 2 (1.3) | 1 (4.0) |
| Spatial Processing Disorder | 46 (6.4) | 10 (5.0) | 2 (1.3) | 0 |
| Mean Number of NDs (StDev) | 3.7 (2.5) | 3 (2.5) | 0.72 (1.7) | 2.5 (2.0) |

Note: Frequencies reflect the number of parents who responded “yes” or “maybe” to their child being ND and having each ND condition.

**Table 7.** Neurodivergent conditions in the autistic CYP and non-autistic ND CYP. For the autistic CYP, these are likely classified as co-occurring conditions. For the non-autistic CYP, these may be single diagnoses or co-occurring conditions. Chi-Square analyses explored between-group differences in prevalence of individual ND conditions between the autistic and non-autistic ND CYP (with the bold text highlighting significant differences). The ‘No SD’ column on the right is for illustrative purposes only and represents the percentage of ND CYP (out of the total number of ND CYP in the sample) that did <u>no</u>t experience SD.

| | Autistic<br>(n=769) | Non-Autistic ND<br>(n=114) | $\chi^2$ | df | n | Sig | No SD |
| --- | --- | --- | --- | --- | --- | --- | --- |
| Autism | 100% (n=769) | 0% | - | - | - | - | 3.25% (n=25) |
| Sensory | **61.6% (n=474)** | **36.8% (n=42)** | **25.648** | **1** | **880** | **< .001** | 1.94% (n=10) |
| ADHD | 58.5% (n=448) | 51.8% (n=59) | 1.841 | 1 | 880 | NS | 2.56% (n=13) |
| Dyslexia | 26.2% (n=201) | 32.5% (n=37) | 1.943 | 1 | 880 | NS | 3.36% (n=8) |
| Dyspraxia | 28.9% (n=221) | 21.9% (n=25) | 2.360 | 1 | 880 | NS | 4.87% (n=12) |
| APD | 18.5% (n=142) | 19.3% (n=22) | 0.038 | 1 | 880 | NS | 3.05% (n=5) |
| Speech Difficulties | 17.6% (n=135) | 10.5% (n=12) | 3.593 | 1 | 880 | NS | 4.75% (n=7) |
| Gifted | 16.2% (n=124) | 17.5% (n=20) | 0.133 | 1 | 880 | NS | 4.16% (n=6) |
| Dyscalculia | 13.2% (n=101) | 8.8% (n=10) | 1.754 | 1 | 880 | NS | 3.6% (n=4) |
| Language Disorder | 12.9% (n=99) | 7.9% (n=9) | 2.331 | 1 | 880 | NS | 3.7% (n=4) |
| Visual Processing Difficulties | 10.6% (n=81) | 6.1% (n=7) | 2.168 | 1 | 880 | NS | 0% |
| Tic Disorder | **10.2% (n=78)** | **1.8% (n=2)** | **8.530** | **1** | **880** | **< .01** | 6.25% (n=5) |
| Unspecified Learning Disorder | 8.5% (n=65) | 6.1% (n=7) | 0.727 | 1 | 880 | NS | 1.39% (n=1) |
| Intellectual Disability | 7.4% (n=57) | 3.5% (n=4) | 2.379 | 1 | 880 | NS | 3.22% (n=2) |
| Other | 12% (n=92) | 16.7% (n=19) | 1.952 | 1 | 880 | NS | 2.6% (n=3) |

Interestingly, Lifelong EHE CYP were equally likely to be described as ND by their parents as CYP in both SD groups [(Current SD=Past SD=Lifelong EHE)>No SD, p<.008 (Bonferroni adjusted alpha level)]. The OR for a CYP in the Lifelong EHE group to be ND (considering “yes” and “maybe” responses) relative to a CYP in the No SD group was 25.8 (95% CI 7.26, 91.46), suggesting the Lifelong EHE CYP had neurodevelopmental profiles comparable to the CYP in the two SD groups.

Co-occurrence between neurodivergencies was high, with many CYP having multiple neurodivergencies [overall mean=3.14 (StDev=2.62); Current SD=3.70 (StDev=2.51); Past SD=3.0 (StDev=2.54), No SD=0.72 (StDev=1.73); Lifelong EHE=2.52 (StDev=2.0)]. Number of ND conditions per CYP differed significantly between the four groups (H(2)=218.123, p<.001), with No SD CYP having significantly fewer ND conditions than all three other groups (all p-values<.001). No significant differences were found between the Current SD and Lifelong EHE groups (p=0.123), or the Lifelong EHE and Past SD groups (p=1.00).

Co-occurrence between neurodivergencies and health difficulties, particularly MH difficulties, was also high. Notably, 89.14% of CYP in the Current SD group whose parents listed one or more health difficulties were also neurodivergent. This figure includes CYP with MH difficulties only, and those with both physical and MH difficulties (see Figure 3). Having a physical health condition only accounted for 0.88% of cases of Current SD CYP, and being both ND and having a physical health condition but no MH difficulties accounted for just 3.5% of cases. Neurotypical CYP currently experiencing SD alongside a MH condition accounted for 6.13% of cases.

**Fig. 3.**
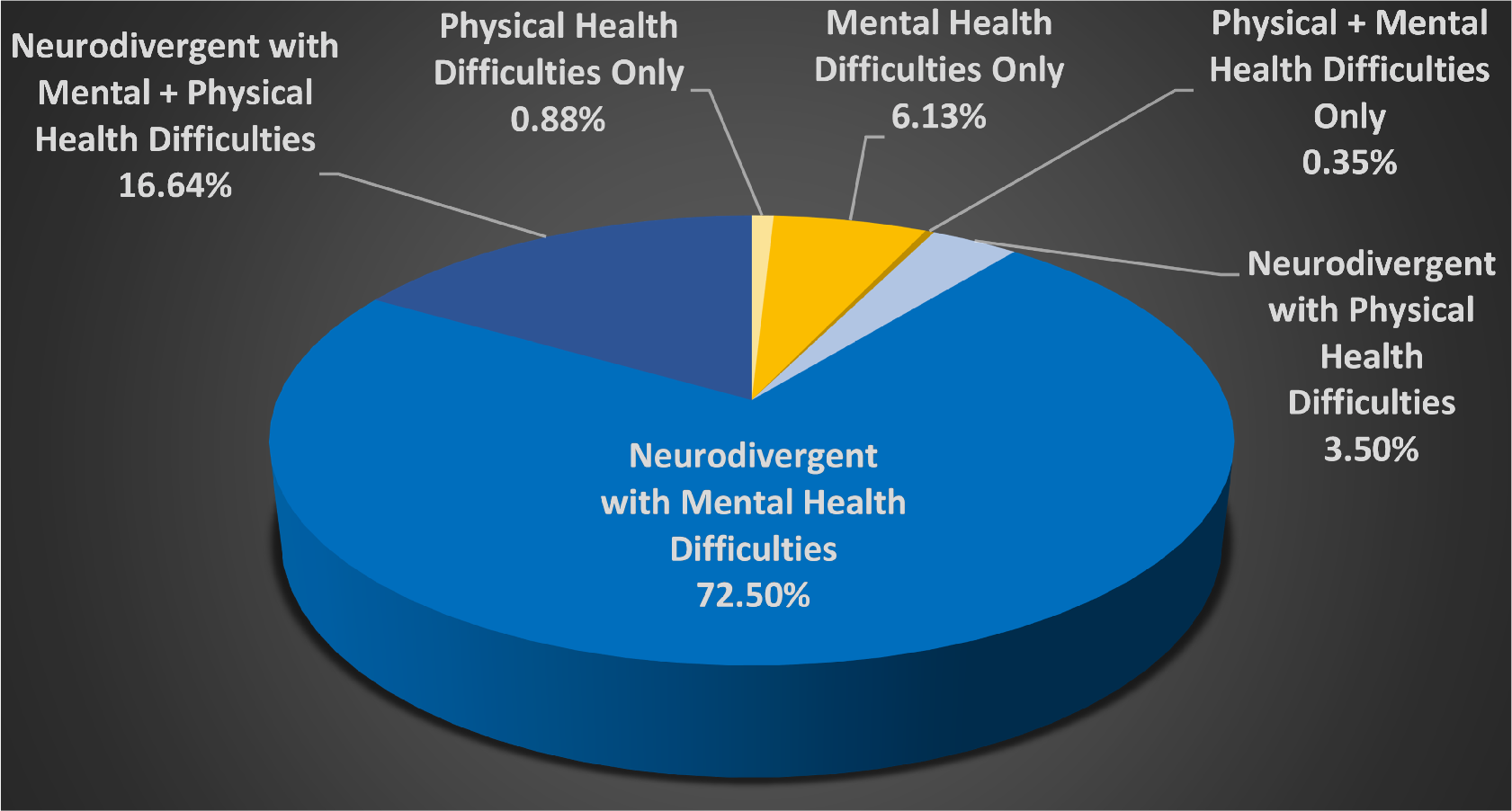
Health Difficulties in the Current SD CYP (as listed by parents in an optional free text box) x ND. Note: ND includes sensory difficulties. Health Difficulties includes the conditions described in Table 4.

#### Autism

Autism was the most prevalent ND condition amongst CYP with SD experience (Current SD: 83.4%; Past SD: 66.2%; see Table 6). These prevalence rates include all individuals who were either diagnosed or suspected to be autistic, and thus prevalence rates were lower when analysis included only CYP with a confirmed autism diagnosis (Current SD: 46.9%; Past SD: 42.1%). Notably, however, this latter method misses 173 (23.4%) Current SD and 20 (9.6%) Past SD CYP who are currently on an autism assessment pathway, 15 (2%) Current SD and 3 (1.4%) Past SD CYP who have had their referral rejected before assessment, and 66 (8.9%) Current SD and 22 (10.5%) Past SD CYP for whom autism is indicated but a referral has not yet been made.

A Chi-Square test revealed a significant difference in the frequency of autism between groups, **χ**²(3,1092)=269.7, p<.001, whereby CYP in both SD groups were more likely to be autistic than No SD CYP. Moreover, Current SD CYP were significantly more likely to be autistic than Past SD CYP (Current SD>Past SD>No SD).

Notably, the OR of an autistic CYP (suspected or diagnosed) experiencing SD (Current or Past) was 37.69 (95% CI [23.22, 61.18]), relative to non-autistic CYP. This increased to 46.61 (95% CI [24.67, 88.07]) when analysis included only autistic CYP with confirmed diagnoses.

Lifelong EHE CYP were also significantly more likely to be autistic than No SD CYP, but were less likely to be autistic than Current SD CYP (Current SD>Lifelong EHE>No SD). The odds of a Lifelong EHE CYP having a confirmed autism diagnosis was significantly greater than for No SD CYP [OR=6.44 (95% CI 0.98, 42.46)]. This increased further when non-diagnosed autistic CYP were included [OR=20.11 (95% CI 5.33, 75.85)]. There was no difference in the prevalence of autism between Past SD and Lifelong EHE CYP.

#### Sensory Processing Difficulties

The second most prevalent ND amongst CYP with SD experience was Sensory Processing Disorder/Sensory Integration Disorder (SPD/SID; see Table 6). The results of a Chi-Square test revealed a significant difference in frequency of SPD/SID across the SD (Past and Current SD combined) and No SD groups [**χ**2(1,1064)=114.372, p<.001]. Visual inspection of Figure 4 (column 2) shows the markedly increased prevalence of SPD/SID in CYP with SD (top panel) relative to those without SD (bottom panel), across the breadth of NDs and differing levels of anxiety and demand avoidance (with only a few exceptions). Notably, CYP within the Lifelong EHE group were also significantly more likely (n=13/25) than CYP without SD (n=5/50) to have SPD/SID (**χ**2(1)=14.501, p<.001; analysis conducted using the Lifelong EHE CYP and their age-matched No SD group).

**Fig. 4.**
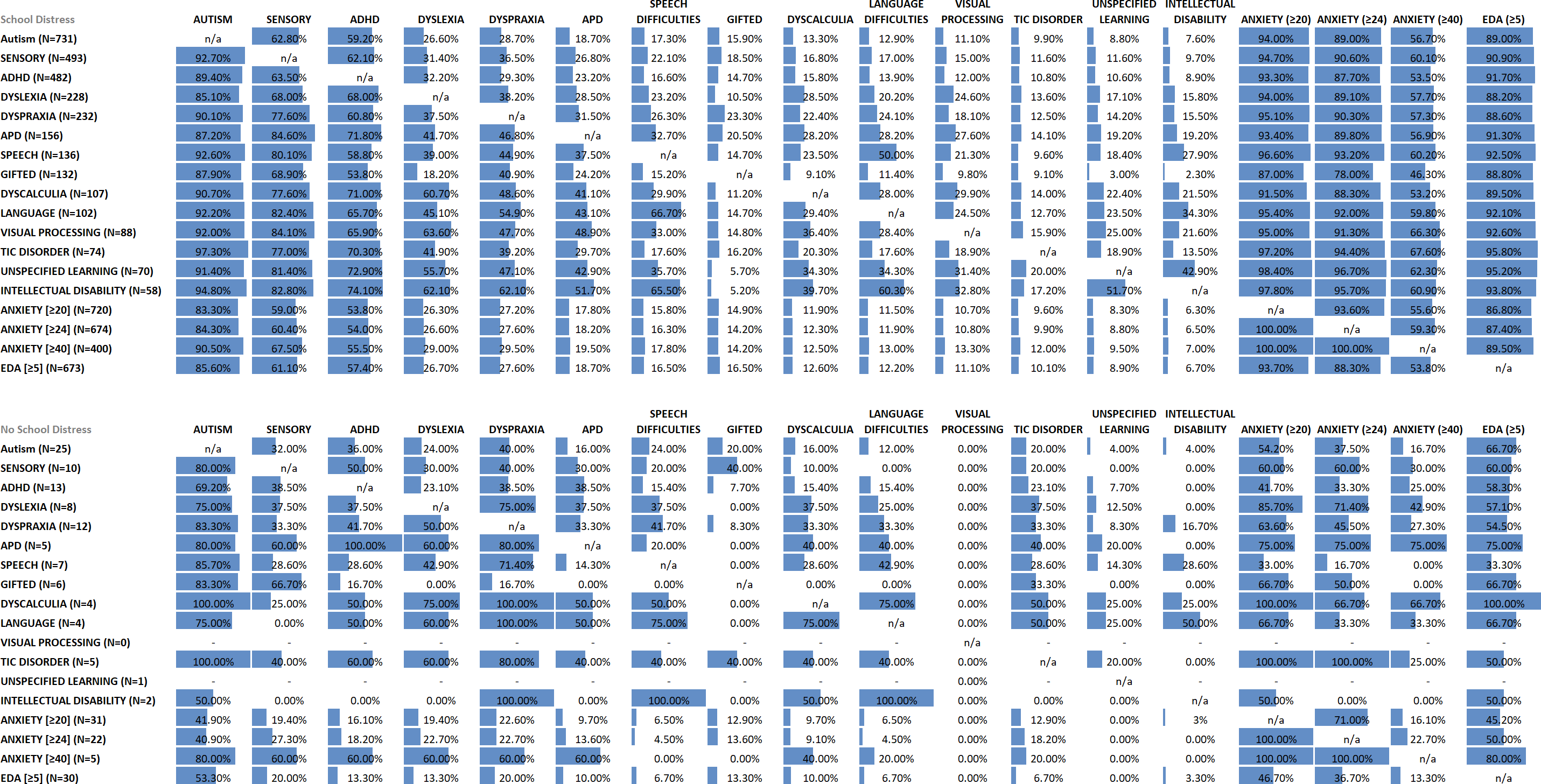
Neurodivergent and anxiety profiles of the CYP with (top panel) and without (bottom panel) School Distress. Each row represents the number of CYP in that group with a specific neurodivergency, or who scores at or above a particular score on the ASC-ASD-P (Anxiety) and EDA-8 (Extreme Demand Avoidance) questionnaires. The columns then indicate the % of these CYP who fall in the other neurodivergencies and Anxiety/EDA groups.

In cases where SPD/SID was indicated, difficulties were reported in an average of 4.8 sensory systems (StDev=2.1). When split by group, the mean number of systems impacted was 4.79 for Current SD CYP (StDev=2.08), 4.96 for Past SD CYP (StDev=2.09), 4.1 for No SD CYP (StDev=2.6) and 4.62 for Lifelong EHE CYP (StDev=2.53). The tactile system, followed closely by the auditory system (both>80%), were the systems identified most frequently as being impacted (see Table S2, upper panel). Having difficulties in just one sensory system was rare, accounting for 3.7% of reported cases.

Across all CYP (including those for whom SPD/SID was not reported), difficulties were reported in an average of 2.28 (StDev=2.8) sensory systems [Current SD=2.72 (StDev=2.8); Past SD=2.19 (StDev=2.8); No SD=0.27 (StDev=1.2); Life-long EHE CYP=2.61 (StDev=3)]. A Kruskal-Wallis test indicated the number of systems impacted differed between groups, H(3)=111.340, p<.001, with pairwise comparisons revealing: Current SD (Mdn=2)>No SD (Mdn=0), p<.001; Past SD (Mdn=0)>No SD, p<.001; and Current SD>Past SD, p=.046.

For CYP experiencing SD, the number of sensory systems impacted (range: 0-8) correlated significantly with SD duration (rs=0.153, p<.001), age of onset of SD (rs=0.214, p<.001), and school attendance in the previous 4 weeks (rs=0.141, p=.002), 2021/22 academic year (rs=0.199, p<.001), and 2020/21 academic year (rs=0.137, p=.003). Number of sensory systems impacted also correlated with anxiety (rs=0.422, p<.001), EDA (rs=0.403, p<.001), and the degree of emotional distress associated with school attendance (rs=0.319, p<.001).

To explore prevalence of SPD/SID further, we subdivided CYP into those who were autistic and those who were non-autistic but otherwise ND. Neurotypical CYP were excluded as they, by definition, did not have any sensory processing differences. 61.6% of the autistic CYP and 36.8% of the non-autistic ND CYP were reported to have SPD/SID. The results of a Chi-Square test found a significant difference in frequency of SPD/SID across the autistic and non-autistic ND groups [**χ**2(1,880)=25.648, p<.001].

Finally, when SPD/SID was indicated, the mean number of sensory systems impacted for autistic CYP (mean=4.89, StDev=2.08, Mdn=5) was significantly greater than for non-autistic ND CYP (mean=3.81, StDev=2.23, Mdn=0, U=56871.5, p<.001).

#### Other Neurodivergent Conditions

The third most prevalent ND amongst individuals with SD experience was ADHD, followed by Dyslexia, Dyspraxia, Auditory Processing Disorder (APD), Speech Difficulties, and Giftedness (see Table 6). The prevalence of intellectual disabilities was relatively low in both SD groups (6.7% Current SD; 5% Past SD). Under the “other” category, hypermobility, PDA, and dysgraphia were the most frequently mentioned NDs.

Given the high co-occurrence between ND conditions and the high proportion of autistic CYP amongst the ND CYP in the sample, there were insufficient cases to contrast prevalence rates for isolated ND conditions.

We did, however, compare prevalence of each ND condition across autistic and non-autistic ND CYP (see Table ??). Other than SPD/SID [autistic group>non-autistic ND group], only Tic Disorders were reported more frequently in autistic CYP (10.2%) than in non-autistic ND CYP (1.8%) (**χ**2(1,880)=8.53, p<.001), and there were no instances where a ND condition was more prevalent in the non-autistic ND group relative to the autistic group.

Breaking this down further to explore co-occurring conditions amongst autistic CYP with and without SD, we found a significantly higher frequency of SPD/SID in the autistic CYP with SD experience [Current and Past SD combined] than in the autistic CYP with no SD experience (**χ**2(1)=9.692, p=.002). Moreover, non-autistic CYP with SD were significantly more likely to have ADHD (**χ**2(1)=29.617, p<.001), APD (**χ**2(1)=11.579, p=.001), Dyscalculia (p=.007, Fisher’s Exact test), Dyslexia (**χ**2(1)=19.998, p<.001), Dyspraxia (**χ**2(1)=11.518, p=.001), Giftedness (**χ**2(1)=8.666, p=.003), SPD/SID (**χ**2=21.627, p<.001), and ‘Other’ ND conditions (**χ**2(1)=9.385, p=.002) than non-autistic CYP without SD (see Figure 5).

**Fig. 5.**
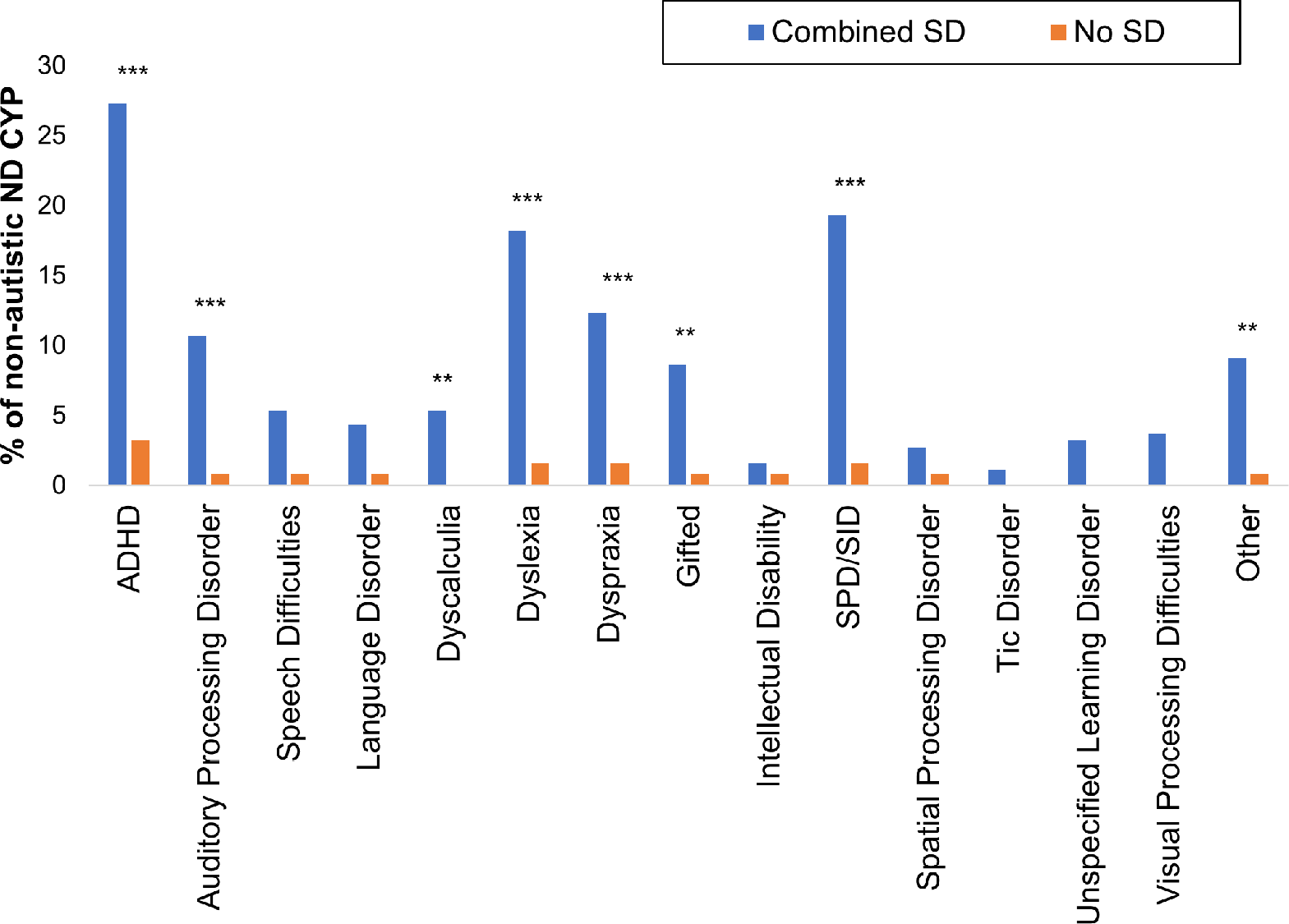
NDs amongst non-autistic CYP with and without School Distress. Note: ‘Combined SD’ includes children in the Current and Past SD groups. * p<.05; **p<.01; ***p<.001.

#### Anxiety

Individual total scores on the ASC-ASD-Parent Version (65) ranged from 0-72.

Only 7.5% of Current SD CYP did not reach the cut-off indicative of significant anxiety (cut off = 20), with 92.5% meeting or exceeding this score (see Figure 6, Panel A). Moreover, 86.7% of Current SD CYP scored above 24 and therefore exceeded the more specific cut-off score, and 53.8% of Current SD CYP scored at least twice the initial cut-off (40+). A ChiSquare test revealed a significant difference in the frequency of <20 and 20+ scorers between the Current and No SD groups, **χ**²(1, 771)=353.661, p<.001. The OR of a CYP experiencing SD if they scored 20+ was 44.015 (95% CI 26.773, 72.362).

**Fig. 6.**
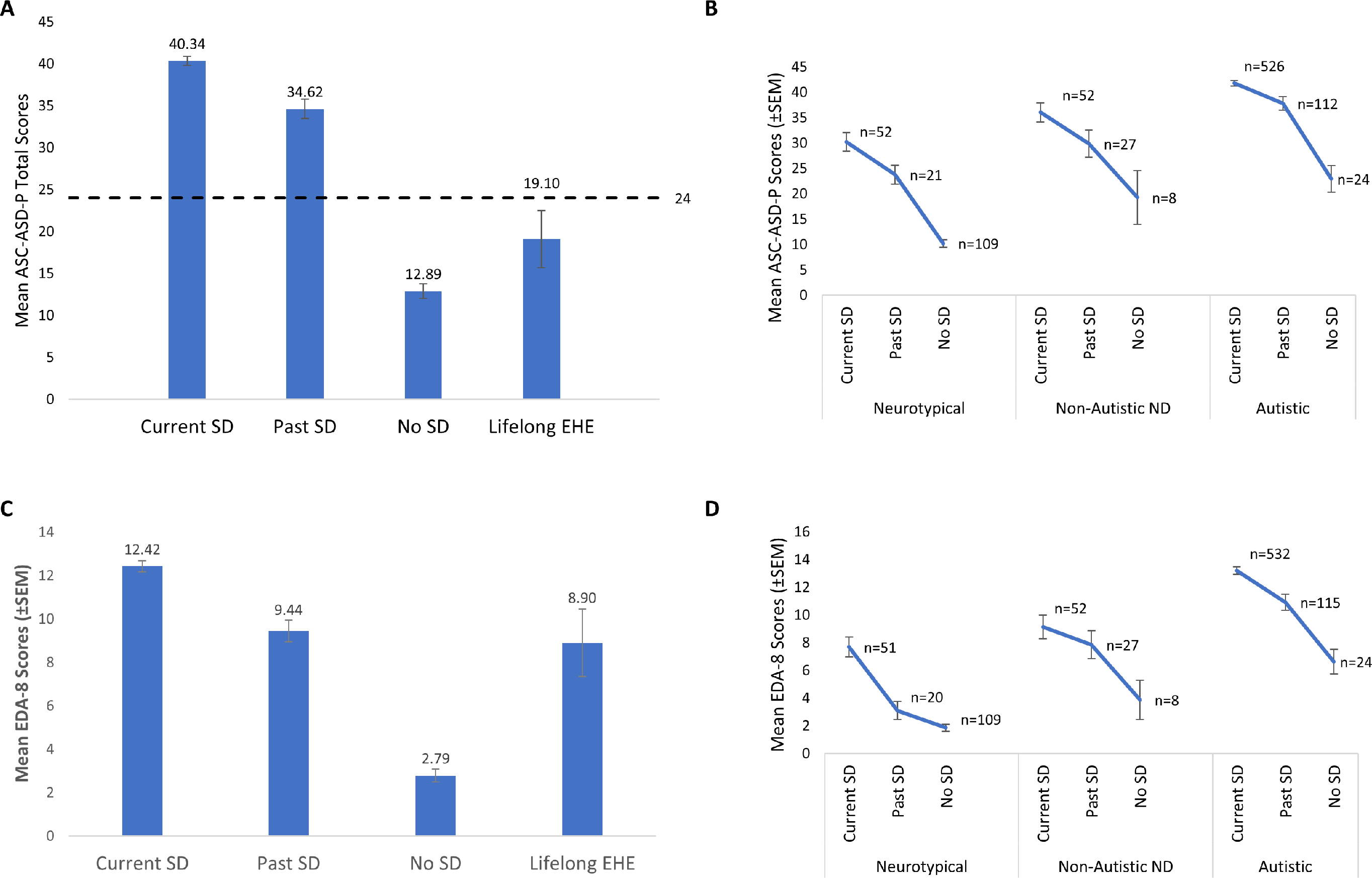
A) Mean total Anxiety scores (ASC-ASD-P) for each of the four groups. The dashed line represents the cut-off score indicative of clinically significant levels of anxiety (i.e., a score of 24). B) Mean total anxiety scores in each group further sub-divided with respect to neurotype. C) Mean Extreme Demand Avoidance scores (EDA-8) for each of the four groups. D) Mean Extreme Demand Avoidance scores (EDA-8) in each group further sub-divided with respect to neurotype. Error bars represent +/1 SEM. Non-Autistic ND = non-autistic, otherwise ND CYP. Note: The Lifelong EHE group are not represented in B and D due to low numbers (e.g., n=2 in the NT group).

Notably, Kruskal-Wallis tests revealed significant between-group differences in anxiety scores, H(3)=296.88, p<.001, with pairwise comparisons indicating No SD CYP (Mdn=10) and Lifelong EHE CYP (Mdn=14.5) had significantly lower total anxiety than Past (Md=33) and Current (Mdn=41) SD CYP [Current SD>Past SD>(No SD=Lifelong SD)]. Additional analyses using the Lifelong EHE age-matched comparison groups revealed Lifelong EHE CYPs’ scores did not differ significantly from those of the No SD CYP, but were significantly lower than those of the Current and Past SD CYP (see Table S3).

We also explored whether anxiety scores correlated with our markers of SD. Higher anxiety correlated significantly with longer SD Duration (rs=0.150, p<.001), more negative impact of school attendance on MH (rs=-0.545, p<.001) and lower school attendance in the previous 20 days (rs=0.41, p=.002), and the 2021/22 (rs=0.199, p<.001) and 2020/21 academic years (rs=0.137, p<.001).

When CYP (n=951) were subdivided into three additional groups (NT, non-autistic ND, autistic), an independent-samples Kruskal Wallis test revealed an overall between-group difference in anxiety (H=260.70, df=2, p<.001). Bonferroni-corrected pairwise comparisons indicated anxiety was significantly lower in the NT group relative to both the non-autistic ND and the autistic group (both p values<.001), and anxiety was significantly higher in the autistic group relative to the non-autistic ND and NT groups (both p values<.001; autistic>non-autistic ND>NT). This pattern was also evident for each SD group individually (see Table 8). The Lifelong EHE group could not be included here due to low numbers in the NT group (n=2).

**Table 8.** Mean and Median Total Scores on the ASC-ASD-P for Autistic, Non-Autistic ND, and Neurotypical CYP in Each of the Four School Distress Groups, and KruskalWallis Tests Investigating Differences in ASC-ASD-P Total Scores Based Upon Neurotype, in Each School Distress Group.

| SD Group | Autistic |  | Non-Autistic ND |  | Neurotypical |  | H(2) | p | Significant group differences |
| --- | --- | --- | --- | --- | --- | --- | --- | --- | --- |
|  | Mean | Median | Mean | Median | Mean | Median |  |  |  |
| Current SD (n=630) | 41.77 | 43.00 | 36.02 | 35.00 | 30.21 | 31.00 | 39.340 | <.001 | Autistic > Non-Autistic ND > Neurotypical |
| Past SD (n=160) | 37.80 | 38.50 | 29.85 | 28.00 | 23.76 | 23.00 | 21.965 | <.001 | Autistic > (Non-Autistic ND = Neurotypical) |
| No SD (n=141) | 22.92 | 20.50 | 19.25 | 17.50 | 10.22 | 8.00 | 24.343 | <.001 | Autistic > Neurotypical;<br>Autistic = Non-Autistic ND;<br>Non-Autistic ND = Neurotypical |
| Lifelong EHE (n=20) | 28.09 | 23.00 | 8.00 | 8.00 | 8.50 | 8.50 | - | - | - |

Note: Kruskal Wallis Tests could not be conducted in the Lifelong EHE group due to low numbers in the neurotypical subgroup (n=2).

This presence of significantly higher anxiety amongst autistic CYP relative to NT and non-autistic ND CYP, regardless of the presence or absence of SD, represents a potential confound when interpreting ASC-ASD-P scores as the significantly higher scores in the SD groups could have been driven by the significantly different rates of autistic CYP between the groups (see above for more detail). It was therefore necessary to compare anxiety levels between the Current, Past, and No SD groups for the NT, non-autistic ND, and autistic groups individually, to ensure anxiety differences persisted when differences in neurodevelopmental profiles were minimised (see Figure 6, Panel B).

Notably, independent-samples Kruskal-Wallis tests found significant between-group differences for anxiety in the NT group (n=182) (H(2)=85.174, p<.001; Current SD=Past SD>No SD), the non-autistic ND group (n=87) (H(2)=10.111, p=.006; Current SD>No SD), and the autistic group (n=662) (H(2)=38.631, p<.001; Current SD>Past SD>No SD). Hence, differences in prevalence rates of autism, and ND more broadly, were insufficient to account for the anxiety differences observed between CYP with and without SD, as these persisted even when neurotype was held constant.

### 5. Demand Avoidance

A Kruskal-Wallis H test revealed significant between-group differences in total EDA-8 scores (H(3)=242.945, p<.001), with post-hoc analyses indicating Current SD CYP had significantly higher scores than Past SD CYP, and Current SD, Past SD, and Lifelong EHE CYP had significantly higher scores than No SD CYP (all p’s<.001; see Figure 6, Panel C). Total scores in the Lifelong EHE group did not differ from those in the Current and Past SD groups, with the latter finding persisting when using the Lifelong EHE age-matched control groups (see Table S3).

Higher EDA-8 scores correlated significantly with longer SD duration (rs=0.095, p=.008), younger age of onset of SD (rs=0.205, p<.001), more negative impact of school attendance on MH (rs=-0.101, p=.011), and worse school attendance in the previous 20 days (rs=0.126, p=.005) and the 2021/22 academic year (rs=0.106, p=.021). Higher EDA-8 scores also correlated significantly with number of sensory systems impacted (rs=0.402, p<.001), and higher anxiety (rs=0.483, p<.001).

Considering autistic relative to non-autistic ND and NT CYP in each SD group separately (see Figure 6, Panel D), independent-samples Kruskal Wallis tests found overall significant between-subgroup differences in EDA-8 scores: 1) Current SD (n=630): H(2)=47.889, p<.001 [autistic>non-autistic ND>NT); 2) Past SD (n=161): H(2)=29.441, p<.001 [autistic>non-autistic ND=NT]; 3) No SD (n=140): H(2)=30.239, p<.001 [autistic>non-autistic ND=NT]; and 4) Lifelong EHE (n=19): H(2)=6.627, p=.036 [autistic>non-autistic ND=NT]. Hence, EDA-8 scores were higher in autistic CYP regardless of SD grouping, suggesting the higher EDA-8 scores in the SD groups may be driven by the higher proportion of autistic CYP in these groups.

However, when exploring each neurotype group in isolation, some significant between-group differences in EDA-8 scores were still evident [NT group (n=180): H(2)=59.009, p<.001, Current SD>Past SD=No SD; non-autistic ND group (n=87): H(2)=6.105, p=.047, no sig. pairwise comparisons; autistic group (n=671): H(2)=34.317, p<.001, Current SD>Past SD>No SD]. Hence, in both the NT and the autistic group, the No SD CYP had significantly lower EDA-8 scores than the Current SD CYP. Thus, differences in rates of autism were insufficient to account for the EDA differences observed between CYP with and without SD, as, alike with anxiety scores, these largely persisted when neurotype was held constant. As with anxiety, the Lifelong EHE group could not be included here due to low numbers in the NT group (n=2).

## Discussion

This study identified several prevalent characteristics amongst CYP affected by SAPs. In most cases, SAPs were underpinned by significant emotional distress associated with school attendance, and parental accounts regarding their child’s difficulties were often harrowing. This led us to devise the term ‘School Distress’ to replace pre-existing terms such as ‘School Refusal’ as this reinforces that, for many CYP, the defining feature of their experience is not a “refusal” to attend school, but rather the severe emotional distress experienced when attempting to do so. Additionally, definitions of ‘School Refusal’ often require CYP to be absent from school for a period of time [e.g., (63, 64) which specify an absence rate of at least 10%-50% in the prior month], thus failing to adequately capture the experiences of many CYP experiencing SD in this study who continued to attend school despite the emotional distress experienced. Given their unaffected attendance rates, and the prior absence of an adequate typology, these CYP’s distress may fall under the radar of educational professionals. Hence, we argue the ‘School Refusal’ label, which captures nothing of the emotional distress suffered by CYP and is deeply unpopular with those with lived experience of SD, should no longer be used. Instead, we propose these difficulties are best described as ’School Distress’, which is not only person-orientated, but also specifically encompasses CYP who manage to attend school despite their distress.

### CYP Characteristics

The CYP with SD experience were young, with onset of their SD commonly occurring within their formative years, and their difficulties were enduring. As hypothesised, SD first occurred significantly earlier and was more enduring in autistic CYP than non-autistic peers, indicating greater SD severity. This replicates and extends previous findings showing that SAPs occur significantly earlier in autistic CYP (20) and aligns with the findings of Munkhaugen et al. (22), whose teacher reports indicated greater severity of SAPs in autistic pupils.

The majority of CYP experiencing SD either currently or previously attended a mainstream provision. Thus, whilst not restricted to mainstream provisions, it appears SD is common in CYP whose educational journey originated in a mainstream setting, posing the question of whether mainstream settings are suitable for all CYP, and if not, which provisions may be more appropriate.

Consistent with the literature, we did not find compelling evidence of differential rates of SD amongst male and female CYP. Notably, 3.3% of parents identified their child as non-binary or transgender, and 1% selected ‘self-describe’, with these options being more frequently selected by parents of CYP with SD experience. Future studies should explore this further to ensure transgender and gender-diverse CYP are being appropriately supported in schools.

Notably, CYP with SD were significantly more likely to be neurodivergent than CYP without SD, confirming our predictions. This is comparable with Epstein et al. (65) who, in a smaller sample, revealed that about 90% of CYP missing school had Special Educational Needs/Disability (SEND) or a health problem. Similarly, 75% of Amundsen et al.’s (27) participants experiencing SAPs were neurodivergent. Notably, co-occurrence of neurodivergencies was high amongst CYP with SD experience, and a large proportion of the CYP experiencing SD were both ND and experienced MH difficulties. The high rate of MH difficulties in our sample is consistent with previous findings showing high levels of depression, anxiety, and stress in CYP experiencing school-related emotional distress [e.g. (41–43)]. However, in this study, neurotypical CYP experiencing SD alongside a MH condition accounted for just 6.13% of cases. Thus, exploring MH difficulties alone [e.g., (42, 43)] is likely to obscure the wider functional profiles of CYP experiencing SD, with our data suggesting such CYP predominantly have multiple neurodivergent conditions alongside MH difficulties.

Autism was the most prevalent ND condition in our sample, with significantly higher rates found amongst CYP with SD experience than without, aligning with previous research. Notably, however, rates of autism here were higher than previous reports [e.g., (20, 22, 27)], with Epstein et al. (65) finding just 40% of the CYP missing school in their sample to be autistic. However, such previous research has typically only measured diagnosed cases of autism, whereas we included CYP whose autism is diagnosed or indicated. Thus, when we restricted our analysis to include only CYP with a confirmed diagnosis, our prevalence rates were more comparable with those in previous research. Unfortunately, such a method misses the large number of CYP who are currently on the assessment pathway, which typically only occurs after considerable evidence of autism has been compiled across settings. This also excludes CYP who have had their referral rejected before assessment, which typically occurs due to services requesting more evidence prior to acceptance, and CYP for whom a referral has not yet been made. Previous research has found no significant differences in autism characteristics between adults with a confirmed diagnosis and those who self-identify as autistic or are awaiting diagnosis (66, 67). Given this, and the very considerable waiting times for an autism assessment in the UK (68), we argue broader inclusion criteria are likely to provide a more accurate estimation of the prevalence of autism amongst CYP with SD.

Notably, SPD/SID was the second most prevalent ND amongst our SD samples and was significantly more prevalent in the SD groups relative to No SD CYP. Additionally, SPD/SID was significantly more prevalent in autistic CYP with SD than in autistic CYP in the No SD group. Finally, although autistic CYP were significantly more likely to have SPD/SID than non-autistic ND CYP, over a third of non-autistic ND CYP were still classified as having SPD/SID. Hence, CYP with SPD/SID, and in particular autistic CYP with SPD/SID, may have a particular vulnerability to SD. Given that co-occurring sensory processing difficulties appear to increase risk of SD in autistic children, this may offer one potential explanation as to why only some autistic children experience SD (22).

To better understand the nature of these difficulties, we explored which and how many sensory systems were affected in CYP experiencing SD. Notably, difficulties within a single sensory system were rare, with CYP with SPD/SID having difficulties across an average of 4.8 sensory systems. Critically, CYP with SD experience had difficulties in significantly more sensory systems than CYP with no SD experience. Moreover, the number of sensory systems impacted correlated significantly with anxiety and all proxy markers of SD, indicating that more pervasive sensory difficulties were associated with more severe SD, as hypothesised. This extends upon past research which highlights sensory difficulties, and the overwhelming sensory demands of the school environment, as reasons as to why CYP can find school distressing (28, 31, 32, 52, 69). Further reinforcing the potential role played by sensory processing difficulties in SD was the observation that just 1.9% of the CYP reported to experience sensory processing difficulties fell into the No SD group. Hence, having no SD was extremely rare amongst the CYP identified by their parent/carer as having SPD/SID.

Interestingly, difficulties were noted in the tactile and auditory systems in 4/5 CYP with SPD/SID. This is notable as tactile hypersensitivity and auditory filtering have previously been linked to cognitive inattention and academic under-performance in autistic CYP in mainstream classrooms (70), potentially providing insight into why individuals with SPD/SID are at increased risk of SD. Relevant also are Howe and Stagg’s findings (71) that autistic pupils attending mainstream school perceived auditory differences to be most disruptive to their learning, followed by touch, smell, and vision. Furthermore, difficulties in the olfactory system were noted in 2/3 CYP with SPD/SID in our study, resonating with observations that “PE changing room” and “incidental smells such as perfume and cleaning products” are particularly challenging sensory experiences for autistic pupils in school (28) (p. 7). Such olfactory processing difficulties, alongside differences in the gustation system (indicated in half of the CYP with SPD/SID reported here), may also explain why many autistic CYP find school halls/canteens particularly distressing. Notably, the sensory difficulties identified in this study align with the findings of Jones et al. (28) who explored the impact of SPD on autistic pupils’ learning and school life, with parental comments including: “They try to protect themselves by covering their ears, closing their eyes, pulling their t-shirts over their noises to block out smells”. In order to fully elucidate the role played by SPD in SD, future studies should seek to further assess the severity of these difficulties, the specific systems in which they occur, and explore these parameters with respect to anxiety, demand avoidance, autism, and ND more broadly.

Given the high co-occurrence of NDs in this study, coupled with autism being so prominent amongst CYP with experience of SD, it was not possible to delineate the individual impact of each ND condition on SD. Despite this, we did find ADHD, APD, Dyscalculia, Dyslexia, Dyspraxia, Giftedness, SPD/SID, and ‘Other’ ND conditions to be significantly more prevalent in non-autistic children with SD experience, compared to those without, indicating that these neurodevelopmental differences may increase risk of SD in the absence of autism. Notably, the high cooccurrence of NDs in our SD sample may be a key finding in itself, whereby it may be the complexity of managing multiple ND conditions within an environment optimised for the NT learner that overwhelms these CYP and renders the school environment so difficult and detrimental to their wellbeing. Support and planning will likely therefore need to be multidimensional and bespoke to the specific needs of individual CYP experiencing SD. Future studies may seek to fully explore the prevalence of SD in CYP with neurodivergent conditions such as ADHD, dyslexia, and dyspraxia, all of which were present at relatively high rates in our SD groups.

The disproportionate rates of ND found amongst SD samples are extremely concerning, and indicate that assessment, diagnosis, and support for ND CYP needs to be improved. Relatedly, care needs to be taken to ensure CYP with SD have access to timely assessments of underlying neurodivergent conditions, with autism, SPD/SID, ADHD, dyslexia, dyspraxia, and EDA (amongst others) all important to consider.

Of additional concern is the finding that over 90% of Current SD CYP met or exceeded the cut-off indicative of significant anxiety symptomatology on the ASC-ASD-P in this study. Moreover, CYP in both SD groups had significantly higher anxiety than CYP who had never experienced SD, regardless of neurotype. When grouped with respect to neurotype and SD experience, the autistic Current SD group had particularly high anxiety scores (mean = 41.52, which is over twice the cut-off score for significant anxiety). Such scores are markedly higher than previously published scores using the ASC-ASD-P in autistic CYP (72). Such elevated scores are of concern, not least because higher anxiety severity is associated with a lower quality of life in both autistic and non-autistic children with anxiety disorders (73), and in autistic CYP more generally (72, 74). Hence, supporting individual CYP experiencing such levels of anxiety should be a priority for educational and health-care professionals. Overall, whilst these findings replicate those of Gonzalvez et al. (42) who also found significantly higher anxiety levels amongst school “refusing” CYP compared to CYP without SAPs, it also extends previous research by using a larger sample size and a broader typology, and considers CYP’s neurodevelopmental profiles. Moreover, it builds on previous research by using a clinical scale devised using evidence of the anxiety phenomenology in autistic CYP specifically, including items relating to sensory anxiety, intolerance of uncertainty, and phobias (75). This is important as anxiety symptoms differ in the context of autism (76), and autistic CYP appear to be at considerably greater risk of SD. Of note, mean anxiety scores in autistic CYP in the No SD and Lifelong EHE groups also exceeded the cut off for significant anxiety, demonstrating the more general heightened anxiety in autistic CYP, however these scores were markedly lower than in the SD groups. Finally, when exploring how anxiety related to our proxy markers of SD, we found higher anxiety significantly correlated with more extensive SD, greater levels of emotional distress due to school attendance, and lower school attendance in the previous 20 days, and the 2020/21 and 2021/22 academic years. Such high anxiety amongst CYP with SD may be a cause or consequence of SD, or both.

As anticipated, CYP with SD experience scored significantly higher on the EDA-8 (62) than CYP in the No SD group. Hence, consistent with parental accounts, CYP with SD appear to display significantly more EDA behaviours than CYP who attend school without difficulty. Additionally, scores on the EDA-8 correlated significantly with all proxy markers of SD severity, as well as higher anxiety, reinforcing the link between EDA and anxiety, and supporting previous anecdotal and research links between Demand Avoidance and SAPs (58, 59). Interestingly, autistic CYP had, on average, higher EDA scores compared to both the NT and non-autistic ND groups, confirming previous accounts linking high levels of demand avoidance to the autistic experience.

Additionally, within this study, scores on the EDA-8 correlated significantly with anxiety scores, and the number of sensory systems in which CYP experience difficulties. These relationships warrant further consideration in order to understand why and how demand avoidant behaviours become so pervasive in autistic CYP (and indeed in some otherwise ND and NT CYP), and how they relate to the emergence and maintenance of SD. This could help address the current dearth of understanding regarding how best to support CYP with demand avoidant profiles in traditional education settings, particularly in mainstream school environments that so often fail to provide inclusive sensory environments for CYP who experience considerable sensory distress (36).

Strikingly, almost one-third of parents in the Current SD group reported that their child received no support at school. Moreover, for many CYP who were on the school’s SEN register (or equivalent) or who had an EHCP, this did not translate into ring-fenced SEN support at school, as indicated by parental comments. Notably, several parents also referred to a lack of support from their child’s school when they attempted to seek additional support, and also mentioned schools blocking their attempts to secure an EHCP for their child. The likely reason for this is well documented elsewhere (i.e., the issues of school budgets and the fact that schools must pay for the first component (£6000) of meeting an EHCP from their own budgets annually (77)).

### Birth order and Sibling SAPs

Within this study we replicated a previous observation in the literature which noted that a high rate of CYP who experience SAPs are the youngest child in their family (78). This was particularly evident in our Current SD group, where almost 40% of CYP were the youngest in their families. Given that we also found that youngest CYP with SD often had an older sibling(s) who had also experienced SAPs (Figure S1), the high rates of youngest children experiencing SD observed here may well be due to multiplier effects, whereby genetic (e.g., ND) and environmental factors (e.g., experience of a previous sibling’s SD or specific SEND provision in their local schools) interact and compound the risk of subsequent children in the family experiencing SD.

### Lifelong EHE CYP and their families

Interestingly, CYP whose parents had decided school-based education was not appropriate for them at an early point in life (Lifelong EHE) showed similar neurodevelopmental profiles to SD CYP, with comparable rates of ND in both the CYP themselves and their family (parents/siblings). Notably, Lifelong EHE CYP were also significantly more likely to have SPD/SID, and to have elevated demand avoidant (EDA) scores, relative to No SD CYP. Importantly, however, there were no significant differences with respect to anxiety scores between the Lifelong EHE CYP and the No SD CYP, and both these groups had significantly lower anxiety scores than the Past and Current SD groups. Thus, despite comparable ND and demand avoidant profiles in the Lifelong EHE and SD CYP, the former group were not experiencing the same levels of anxiety as the CYP who experienced SD (be that currently or historically). Relatedly, whilst 92.2% of Current SD and 84.5% of Past SD CYP met the clinical cut-off indicative of significant anxiety on the ASC-ASD-P, only 40% of Lifelong EHE CYP met this score. Hence, Lifelong EHE CYP, and particularly ND Lifelong EHE CYP, are an important comparison group here, and suggest that ND profile alone is not sufficient to account for these markedly different anxiety profiles. Future research should attempt to explore the emergence of anxiety in both school- and home-educated ND/demand avoidant CYP longitudinally, to understand the drivers of this anxiety more comprehensively.

Of note here is that some of the parents of Lifelong EHE CYP highlighted that their child’s neurodevelopmental profile was a key determinant in their decision to not enrol their child in a school setting. These parents noted recognising early in their child’s life that, given their child’s ND profile, they would likely face difficulties accessing school-based education (see Table S1, Q5). Moreover, some reported that they considered EHE a better fit to their child’s needs as it affords them flexibility to readily adapt their approach to meet their child’s individual learning needs (see Table S1, Q6), or to provide the levels of individual support required (see Table S1, Q7). This flexibility attunes with the advice of specialist PDA educators (61). However, home education is simply not a feasible educational option for many families, not least due to the considerable financial implications. Moreover, on a societal level, is it acceptable for education outside of the family to be inaccessible to some CYP, simply as a consequence of ND profile?

## Limitations

One key limitation of this study is the lack of diversity amongst our sample, with an over-representation of white CYP, meaning our findings may not accurately represent the profiles of all CYP experiencing SD. Our Current and No SD groups were also living in less deprived areas than would be expected by chance alone. Thus, future research should aim to collect more diverse samples to gain a more comprehensive understanding of the experience of SD across all CYP, and to therefore guide more individualised support. Moreover, this study was limited to the UK, further reducing the generalisability of our findings. As education systems vary internationally, the SD experience and the characteristics of individuals experiencing SD may differ between countries, providing an additional avenue for future research. Additionally, whilst this study was advertised widely on social media, the sites where it was shared may have influenced who participated. For example, within our No SD group, 16.8% of CYP were autistic, despite the national prevalence rates of autism standing at around 1-2% (21), indicating this group may not be entirely representative of all CYP who do not experience SD. Within this study, we used a broad criterion for ND conditions, including CYP currently awaiting assessment/diagnosis, CYP whose referral was rejected, and CYP who have yet to be referred. The rationale behind this is discussed at length above, however it is possible this led to an over-estimation of prevalence rates. A final weakness is the differing sample sizes between participant groups, with the Lifelong EHE group being particularly small, potentially influencing the accuracy of between-group comparisons. Future research should therefore try to collect more evenly-sized participant groups, although this is challenging in rarer groups such as Lifelong EHE CYP. This disparity in sample size did, however, arise due to the volume of CYP currently experiencing SD in the UK.

One key drawback of the SD literature generally, as opposed to this study specifically, is the lack of a standardised questionnaire to assess SD which is suitable for use in autistic individuals. Given the prevalence of autism amongst CYP experiencing SD, development of such a questionnaire which can be used in clinical, education, and research settings is vital. Thus, one next step should be to gather perspectives of autistic CYP and autistic adults, and to work collaboratively with them to develop a standardised questionnaire to assess SD severity and/or risk.

This study also had several strengths, including the large number of participants who were parents of CYP experiencing SD, and the exploration of various aspects of SD within this large sample size. This was much greater than in previous research, enabling stronger conclusions to be made.

## Conclusions

The Human Rights Act states “No person shall be denied a right to an education” (26). However, our findings suggest that large numbers of CYP and, in particular, neurodivergent CYP are currently unable to access education within the United Kingdom. As such, additional research involving CYP who experience SD, and their families, is urgently needed to further understand this problem, and to develop solutions which ensure all CYP are able to access education, and which ensure that all families have the option to avail of safe educational opportunities for their child outside of the family. Such research should be used to inform future changes to education legislation, as legislation changes which do not consider this evidence-base run the risk of making the current situation even worse for those CYP at risk of distress-based school attendance difficulties.

Wider discussion with respect to the appropriateness of traditional, school-based education for all CYP is also needed. Moreover, further research, ideally co-produced with autistic and otherwise ND individuals, is needed to determine best practices in education, and to ensure appropriate understanding of how ND pupils learn best (57). Relatedly, research into best-pedagogical practice for pupils with SEND, including pupils with complex presentations, is urgently needed, especially within mainstream settings (79). Given the substantial heterogeneity in the neurobiology of autism, this will undoubtedly be complex, and efforts must include consideration of how learning needs will vary with neurosubtype (80, 81) and demand avoidant profile (56). Research exploring educational and life outcomes of Lifelong EHE CYP, and CYP with provisions such as EOTAS, is also urgently required to better understand how CYP can be successfully educated outside of school settings.

## Data Availability

All data produced in the present study are available upon reasonable request to the authors following the publication of the full dataset.

## ACKNOWLEDGEMENTS

We sincerely thank all the parents who generously gave up their time to take part in this research. We wish them, and their children, all the very best in the future. We also thank Katherine Patterson and Eliza Hockey for their assistance with initial data processing and analysis, and Kieran Rose for his advice on language.

## Supplementary Note 1: School Distress Siblings

**Fig. S1.**
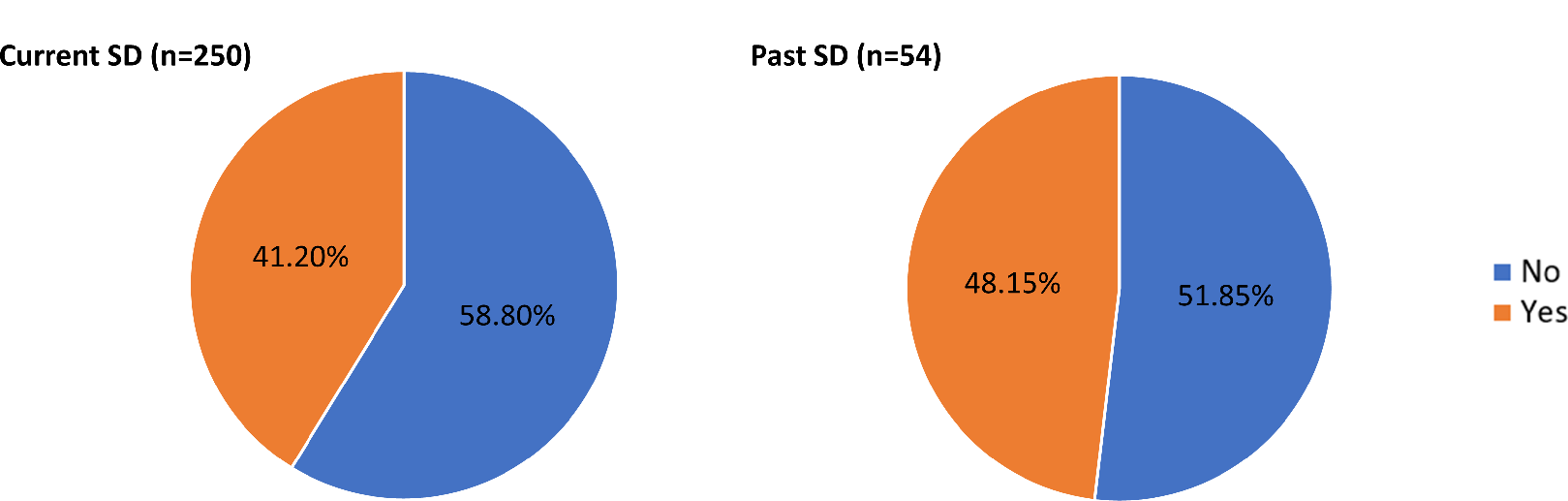
Presence of School Attendance Difficulties in Any of the Siblings of CYP in the Current and Past SD Groups Who Were the Youngest Child in Their Family.

## Supplementary Note 2: Additional Quotes Provided by Parents

**Table S1.**
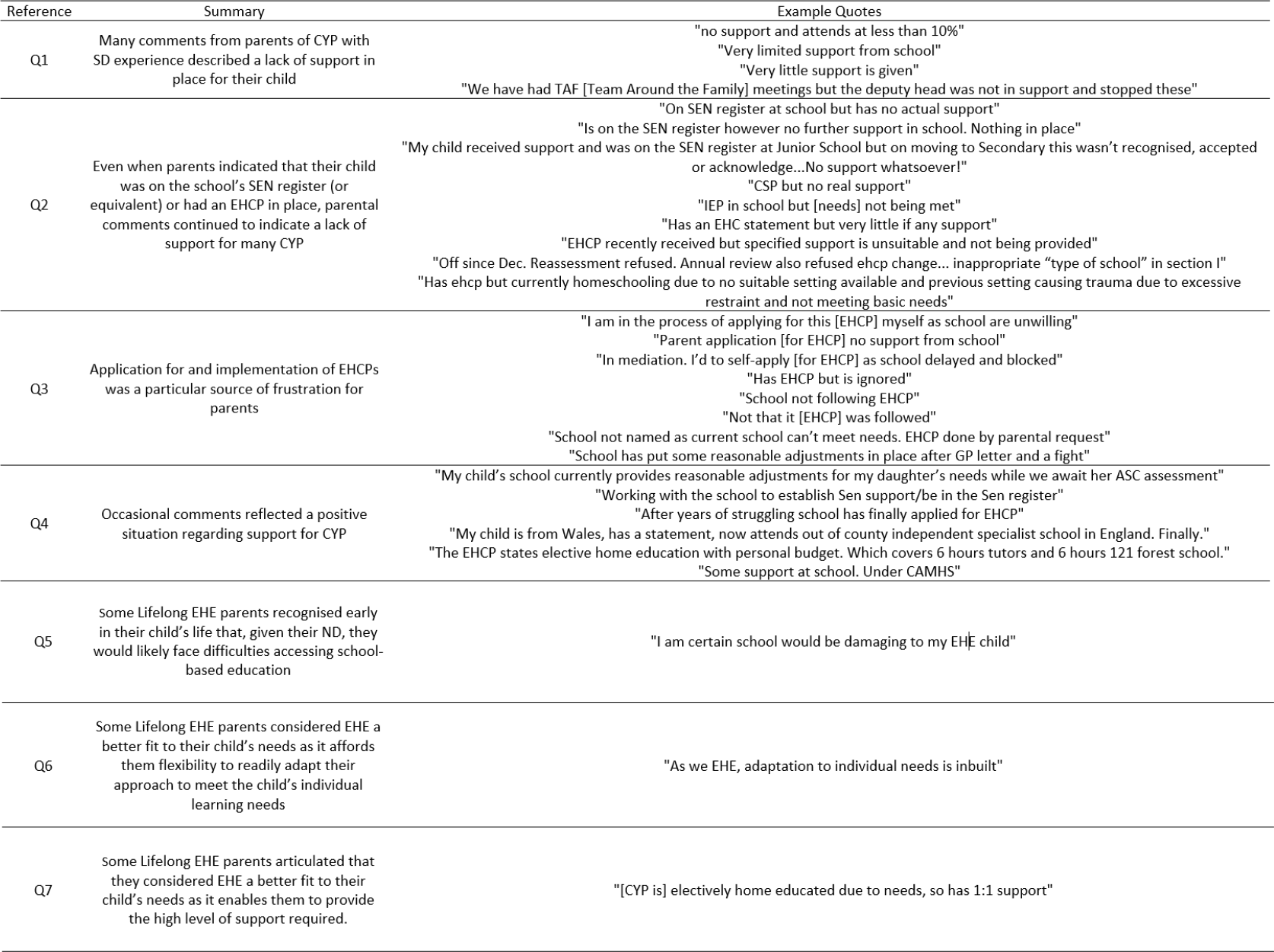
Additional Quotes Provided by Parents Throughout the Survey in Response to Various Questions.

## Supplementary Note 3: Sensory Processing Difficulties: Additional Information

**Table S2.** Sensory Processing Difficulties: Sensory Systems Affected and Mean Number of Sensory Systems Affected.

| Sensory System | Current SD | Past SD | No SD | Lifelong EHE |
| --- | --- | --- | --- | --- |
| <u>CYP with SPD</u> |  |  |  |  |
| Tactile | 84.6% | 87.1% | 80.0% | 92.3% |
| Auditory | 82.6% | 81.2% | 90.0% | 84.6% |
| Interoceptive | 67.4% | 60.0% | 70.0% | 61.5% |
| Olfactory | 67.4% | 60.0% | 70.0% | 61.5% |
| Proprioceptive | 52.0% | 63.5% | 50.0% | 69.2% |
| Gustation | 51.7% | 55.3% | 30.0% | 53.8% |
| Visual | 41.3% | 40.0% | 20.0% | 23.1% |
| Vestibular | 38.8% | 42.4% | 30.0% | 30.8% |
| <u>All CYP</u> |  |  |  |  |
| Tactile | 48.1% | 38.3% | 5.4% | 52.2% |
| Auditory | 47.0% | 35.8% | 6.0% | 47.8% |
| Interoceptive | 38.3% | 26.4% | 4.7% | 34.8% |
| Olfactory | 34.5% | 29.5% | 2.7% | 26.1% |
| Proprioceptive | 29.6% | 28.0% | 3.4% | 39.1% |
| Gustation | 29.4% | 24.4% | 2.0% | 30.4% |
| Visual | 23.5% | 17.6% | 1.3% | 13% |
| Vestibular | 22.1% | 18.7% | 2.0% | 17.4% |

## Supplementary Note 4: Statistics: ASC-ASD-P and EDA-8

**Table S3.** Medians, IQR, and Kruskal-Wallis Tests Investigating Differences in ASC-ASD-P and EDA-8 Total Scores Between Groups Age-Matched to the 20 Lifelong EHE CYP for Whom Data was Available.

| Measure | Current SD |  | Past SD |  | No SD |  | Lifelong EHE |  | H(3) | p | Significant group differences* |
| --- | --- | --- | --- | --- | --- | --- | --- | --- | --- | --- | --- |
|  | Median | IQR | Median | IQR | Median | IQR | Median | IQR |  |  |  |
| ASC-ASD-P Total | 41 | 23.5 | 30.5 | 18.8 | 12 | 9 | 16 | 22 | 61.25 | <.001 | Current SD = Past SD > No SD = Lifelong EHE |
| EDA-8 Total | 14 | 10.8 | 9 | 10 | 2 | 5 | 7 | 8 | 45.68 | <.001 | Current SD = Past SD > No SD, Past SD = Lifelong EHE, Current SD > Lifelong EHE, Lifelong EHE > No SD |

IQR = Interquartile Range.
\*Pairwise comparison. Significance values have been adjusted by the Bonferroni correction for multiple tests.

